# Tranexamic acid in reducing expected blood loss in moderate to low risk surgeries: systematic review, meta-analysis and cost effectiveness analysis

**DOI:** 10.1101/2025.07.21.25331903

**Authors:** Nishant Jaiswal, Giorgio Ciminata, Will Robinson, Martin Taylor-Rowan, Tom Morris, Clareece Nevill, Hadiqa Tahir, Elizabeth Fisher, Ryan Mulholland, Michael Lumsden, Anna Noel-Storr, Andrew Davies, Nicola J Cooper, Terry Quinn, Alex Sutton, Olivia Wu

## Abstract

Tranexamic acid (TXA) is well-established as a safe intervention for reducing transfusion requirements in surgeries with high-risk for blood loss. However, its role in surgeries classified as low-risk for blood loss remains uncertain. Given the frequency of such procedures, even small clinical benefits could have substantial cumulative impact. We assessed the clinical and cost-effectiveness of TXA in surgeries with low expected blood loss A systematic review and meta-analysis of randomised controlled trials (RCTs) for adults or children undergoing low-risk surgeries, comparing peri-operative TXA (any route or dose) with placebo or standard care informed the clinical effectiveness and a decision model adapted from NICE NG24, focusing on short-term hospital costs informed the cost-effectiveness analysis. We included 82 RCTs comprising 8506 participants. TXA significantly reduced blood loss (ratio of means 0·73, 95% CI 0·68,0·79) and transfusion rates (odds ratio 0·39, 95% CI 0·25,0·61). It also reduced hospital stay by 0·4 days (MD = -0·40 days, 95% CI = -0·77, -0·02) and improved pain scores at 1 and 2 weeks postoperatively. Evidence for thrombotic events was limited and inconclusive. The cost-effectiveness analysis showed TXA was cost-saving (£156 per patient) and had a 99% probability of being cost-effective at the £20lJ000 per QALY threshold. Reduction in bleeding and improved recovery outcomes even in surgeries with low anticipated blood loss support broader use of TXA in surgical care and suggests revisiting existing guidelines to include surgeries with any bleeding risk. Further research should examine long-term safety and patient-reported outcomes.

## Introduction

Peri-operative bleeding remains a major risk leading to adverse outcomes and increased healthcare resource use and costs, this is despite advances in surgery, anaesthesia and transfusion medicine.^1^ Allogeneic blood transfusion, standard for surgical blood loss, can lead to direct and indirect complications including transfusion reactions, infection, and immunomodulation.^2,3^ Additionally, the availability of blood products is a growing global concern.^4^ Thus, issues with both safety and availability highlight the need for alternative strategies to manage perioperative bleeding and reduce blood transfusion. Antifibrinolytic agents, such as tranexamic acid (TXA), are proven to reduce blood transfusions following major surgery and have been recommended in surgeries where moderate-to-high blood loss is expected.^5,6^ However, the anticipated blood loss of many common surgeries are less than 500 ml. While this is considered low-risk, the actual volume of blood loss can vary, and some patients in this group may experience considerably higher levels of bleeding than expected. While there is emerging evidence on the effectiveness of TXA in specific surgeries, the overall risk-benefit profile of TXA across surgical specialities classified as low-risk for blood loss has not been comprehensively evaluated. Our aim was to addresses this important gap by collating existing evidence and evaluating the clinical and cost effectiveness of TXA in patients undergoing surgeries classified as low-risk of blood loss.

## Methods

### Clinical Effectiveness of TXA in Low Risk for Blood-Loss Surgeries

We conducted a systematic review and meta-analysis according to the PRISMA 2020 reporting guidelines^7^ and registered the protocol prospectively at PROSPERO with registration number CRD42023467639^8^ (deviations are mentioned in appendix 6).

### Search Strategy and Study Selection

We used complementary approaches to collating evidence following a search strategy developed by our information specialist (ANS) using Cochrane validated randomised controlled trials (RCT) sensitivity-maximising methodological filters where appropriate.^9^ We conducted primary electronic literature searches to identify studies published from 2015 onwards (previous guidelines recommending TXA to reduce blood loss were published in 2015^5^) on databases including: MEDLINE (OVID), Embase (OVID), and Cochrane Library, including Central Register of Controlled Trials to identify RCTs (Appendix 1, table 1). The searches were current as of July 2024. Because our searches focussed on studies published after 2015, we used relevant systematic reviews (identified from the same searches) to find studies published before 2015. Additionally, we searched trial registeries including clinicaltrials.gov and International Clinical Trials Registry Platform (ICTRP) to identify ongoing trials.

**Table 1:** Inclusion criteria for the main and exploratory analysis.

| **Surgeries Classified as Low Risk for Blood Loss** |  |
| --- | --- |
| Participants (P) | Individuals of any age group, gender undergoing elective or emergency surgery classified as low risk for blood loss according to the ISTH criteria except those undergoing caesarean section |
| Intervention (I) | Tranexamic acid administered peri-operatively in any dose or route |
| Comparator (C) | Usual care, no TXA or a placebo |
| Outcomes (O) | Total Blood Loss (TBL),<br>Blood transfusion,<br>Deep vein thrombosis (DVT)<br>Change in Post-operative Pain scores,<br>Length of stay in Hospital |
| Study Type | Randomised controlled trials |
| **Exploratory Analysis** |  |
| Participants (P) | Individuals of any age group, gender undergoing elective or emergency orthopaedic surgeries across high to low risk for blood loss |
| Intervention (I) | Tranexamic acid administered peri-operatively in any dose or route |
| Comparator (C) | Usual care, no TXA or a placebo |
| Outcomes (O) | Total Blood Loss (TBL),<br>Blood transfusion |
| Study Type | Randomised controlled trials |

We formulated the review question in accordance with the PICOS (population, intervention, comparator, outcomes and study type) structure (Table 1). In addition, we included RCTs spanning from high- to low-risk for bleeding from the orthopaedic speciality, for a parallel exploratory analysis to help place the data from low bleeding risk surgery into context (Table 1).

Four reviewers (GC, WR, RM, MTR) screened the titles and abstracts, conducted full text screenings and recorded the reasons for exclusion. The reviewers also conducted 20% duplicate screening. We planned to perform 100% duplicate screening if disagreement between reviewer were greater than 20%.

### Data Extraction and Management

We subdivided the studies by surgical specialities, according to the Royal College of Surgeons of England classification^10^, and risk of bleeding (high, moderate and low) based on the risk stratification for procedural bleed risk as suggested by the International Society on Thrombosis and Haemostasis (ISTH) guidance statement.^11^

Eight reviewers (WR, GC, NJ, HT, CN, RM, EF and ML) extracted data from the included sources using a pre-designed and pre-piloted extraction form using Microsoft Forms (Microsoft Corporation, US). This included the following study characteristics: participants, trial inclusion and exclusion criteria, details of intervention including routes and dosage, comparator details, and outcomes as mentioned in Table 1. Systematic reviews were used to extract data where possible,^12^ including data on risk of bias assessments; however,original trial reports were consulted for additional information when required.

### Risk of Bias Assessment

When needed, one of the three reviewers (NJ, GC, HT) assessed risk of bias using the Cochrane risk of bias tool (RoB 1)^13^ (Appendix 2, Table 1). To ensure high accuracy, another review author (MTR) independently duplicated 10% of the risk of bias assessments.

### Data Synthesis

The clinical effectiveness synthesis was divided into: (i) a primary analysis of trials involving surgeries at low-risk of bleeding, and (ii) an exploratory analysis of orthopaedic trials spanning procedures with high to low bleeding risk.

i. For trials at low-risk of bleeding, we conducted a pairwise meta-analysis using a random-effects model to compare TXA given via any route with the control arm (placebo, or standard care) using the metafor package^14^ in R version 4.3.1.^15^ For multi-arm trials, data from all eligible TXA arms were combined and compared with the control arm, while non-TXA arms were excluded.^16^ For the dichotomous outcomes (participants requiring transfusion and DVT), we reported odds ratios (OR) with 95% Confidence intervals (CIs) and for continuous outcomes (length of hospital stay and pain) we reported mean differences (MD) with 95% CIs. For the primary outcome of total blood volume loss (TBL) during the peri-operative period, we reported the ratio of means (RoM) with 95% CIs.^17^ Median and IQR data were converted to mean and standard deviation (SD) where necessary.^18^
ii. For the exploratory analysis, we used meta-regression to further understand the relationship between mean TBL in the control group (i.e. including ‘baseline risk’ as a covariate) and transfusion odds and TBL outcomes. We performed the analysis initially on the orthopaedics high-risk of bleeding trials and then on the all orthopaedics (i.e. high- and low-risk trials).

### Heterogeneity

We reported between-study heterogeneity using the between-study variance estimate and its associated prediction intervals^19^, alongside the *I*^2^ statistic.^20^ Additionally, we conducted meta-regression and subgroup analyses on surgical speciality and route of TXA administration.

### Publication Bias

We constructed contour-enhanced funnel plots^21^ and regression-adjusted funnel plots removing the impact of covariates to investigate publication bias and the effect of small studies on the primary outcomes.

### Subgroup and Sensitivity Analysis

We performed subgroup analyses for surgery speciality and route of TXA administration. We had also pre-specified subgroup analysis for adult and paediatric populations, but due to lack of uniform reporting of outcomes this subgroup analysis was not feasible. We conducted sensitivity analysis by repeating the meta-analyses using a fixed effect model. Initially, for the length of stay outcome, one trial was excluded due to a SD of 0 for both trial arms (calculated from medians and IQRs); however, this study was included in a sensitivity analysis by assumingSDs of0·1.

### Certainty of Evidence

The Grades of Recommendation, Assessment, Development and Evaluation (GRADE) approach was used to assess the overall certainty of evidence for each primary outcome.^22^ GRADEpro GDT software^23^ was used to construct the summary of findings table.

### Interest-Holder Review

A multidisciplinary group of clinicians from haematology, ear, nose and throat (ENT) surgery, general surgery and orthopaediacs, as well as public contributors, reviewed the findings and provided feedback.

### Cost-Effectiveness Analysis

We conducted a cost-utility analysis to compare the cost-effectiveness of TXA versus standard care for surgical patients at lower risk of bleeding (<500 ml) based on evidence from the clinical review. The structure of the decision model was adapted from that developed and reported in NICE Guideline NG24 for surgeries with high to moderate bleeding risk, with two notable modeifications. Firstly, 30-day mortality was excluded due to lack of evidence for either mortality per se, or TXA’s effect on mortality in low-risk patients. Our focus was therefore on hospital costs and outcomes, without extending to beyond the surgical admission. Secondly, while NG24 modelled the volume of blood required for each transfusion, we assumed one unit of blood per transfusion in low-risk surgeries. The key model inputs were therefore the risk of transfusion in the absence of TXA, and its effect in averting transfusions and reducing hospital stay, each of which was informed by the clincial review.

The cost-effectiveness evaluation was conducted from the perspective of the UK National Health Service (NHS), capturing only costs and outcomes associated with the index hospital admission.^11^ We implemented the model probabilistically (10,000 iterations), reflecting uncertainty quantified by the clincial review, and reported expected net monetary benefits at a cost-effectiveness threshold of £20,000 per QALY. We also performed individual sensitivity analyses for key parameters (Appendix 5).

## Results

The database searches returned a total of 1951 records, of which full texts were retrieved for 1272 records. This resulted in identification of 40 eligible trials and 19 systematic reviews of TXA trials published since 2015.^24-41^ The latter identified a further 42 eligible trials (Figure 1). Thus, in total, 82 RCTs (n=8506 adult and paediatric participants) on surgeries classified as low-risk for blood loss were included in the review (Table 2). Of these 82 included studies, one trial^42^ was subsequently excluded from the blood loss outcome meta-analysis (the only review outcome it reported data on) because of reporting implausibly small standard errors. The risk of bias assessment revealed a substantial proportion of studies at a low risk of bias for all key domains suggesting adherence to core methodological quality. There were ten studies^43-52^ at high risk of bias for at least one domain, particularly domains related to blinding of participant and personnels and outcome assessors, and forty-one studies^42,45,46,49,50,53-90^ at unclear risk of bias for at least one domain, most commonly for random sequence generation and allocation concealment suggesting inadequate information about these domains. Overall, approximately 12% of the total studies were at high risk of bias for any domain thereby suggesting the evidence base was of relatively high quality (appendix 2, Table 2).

**Figure 1:**
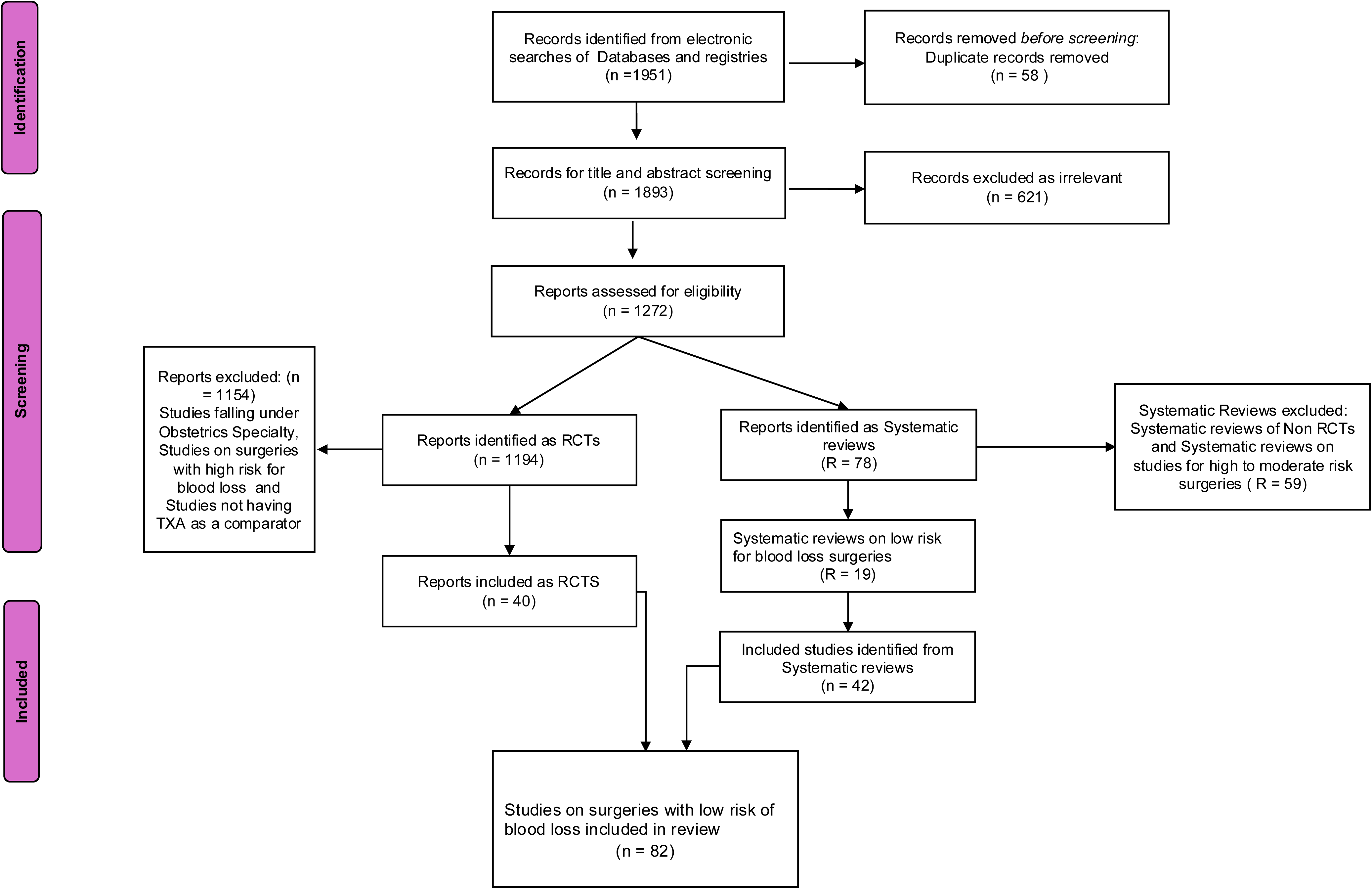
PRISMA flow diagram depicting the inclusion of studies (surgeries classified as low-risk for blood loss)

**Table 2:**
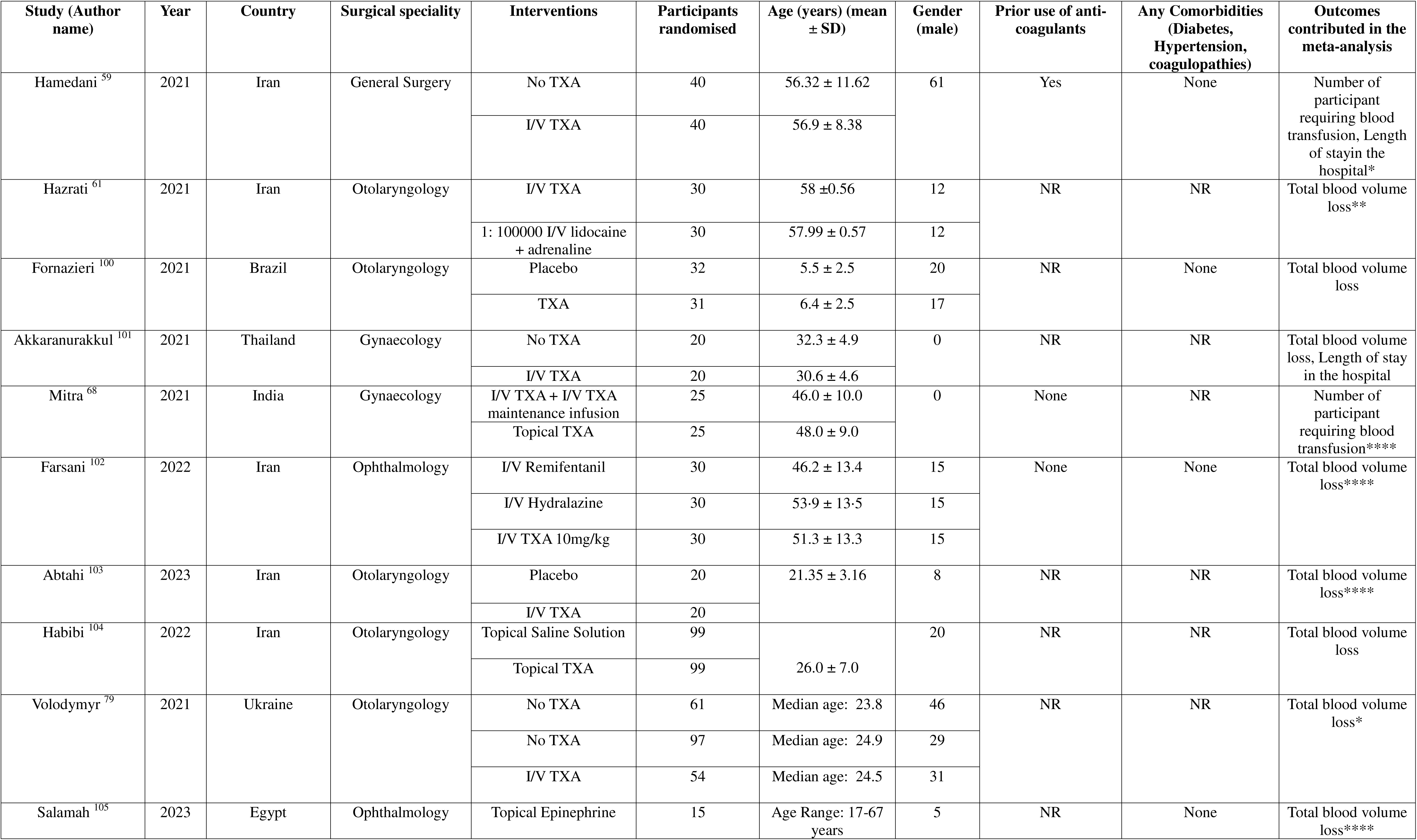
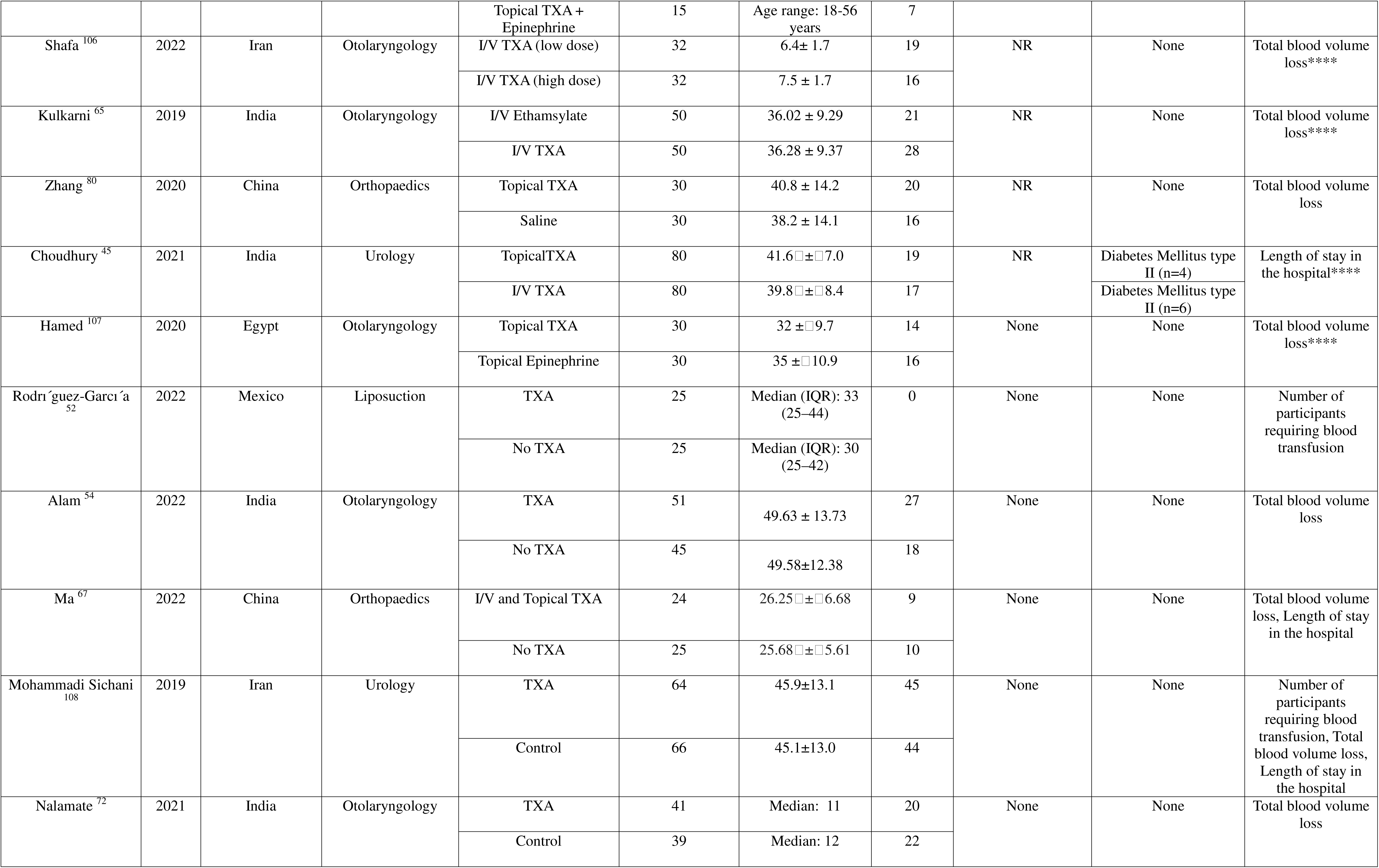
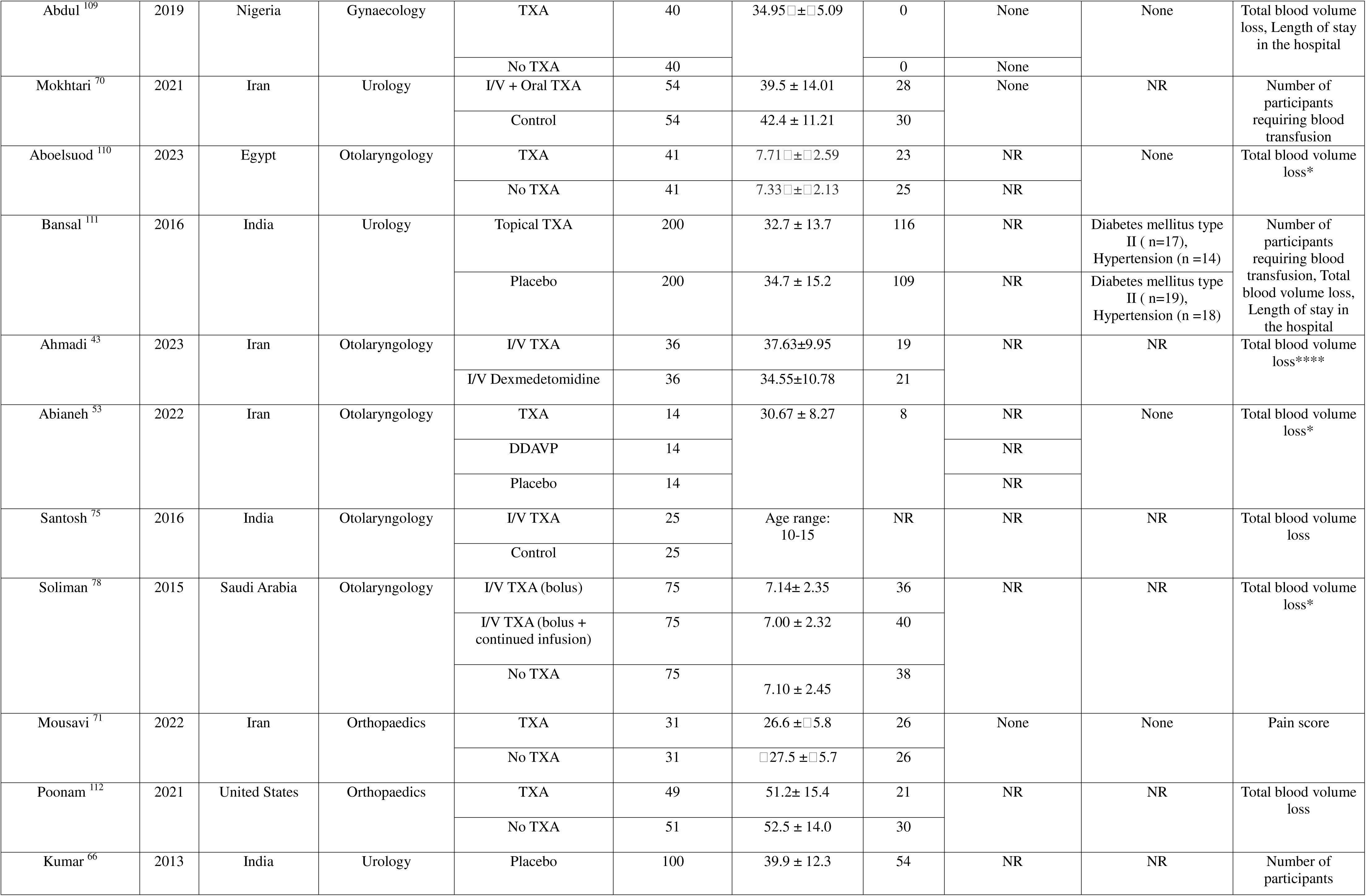
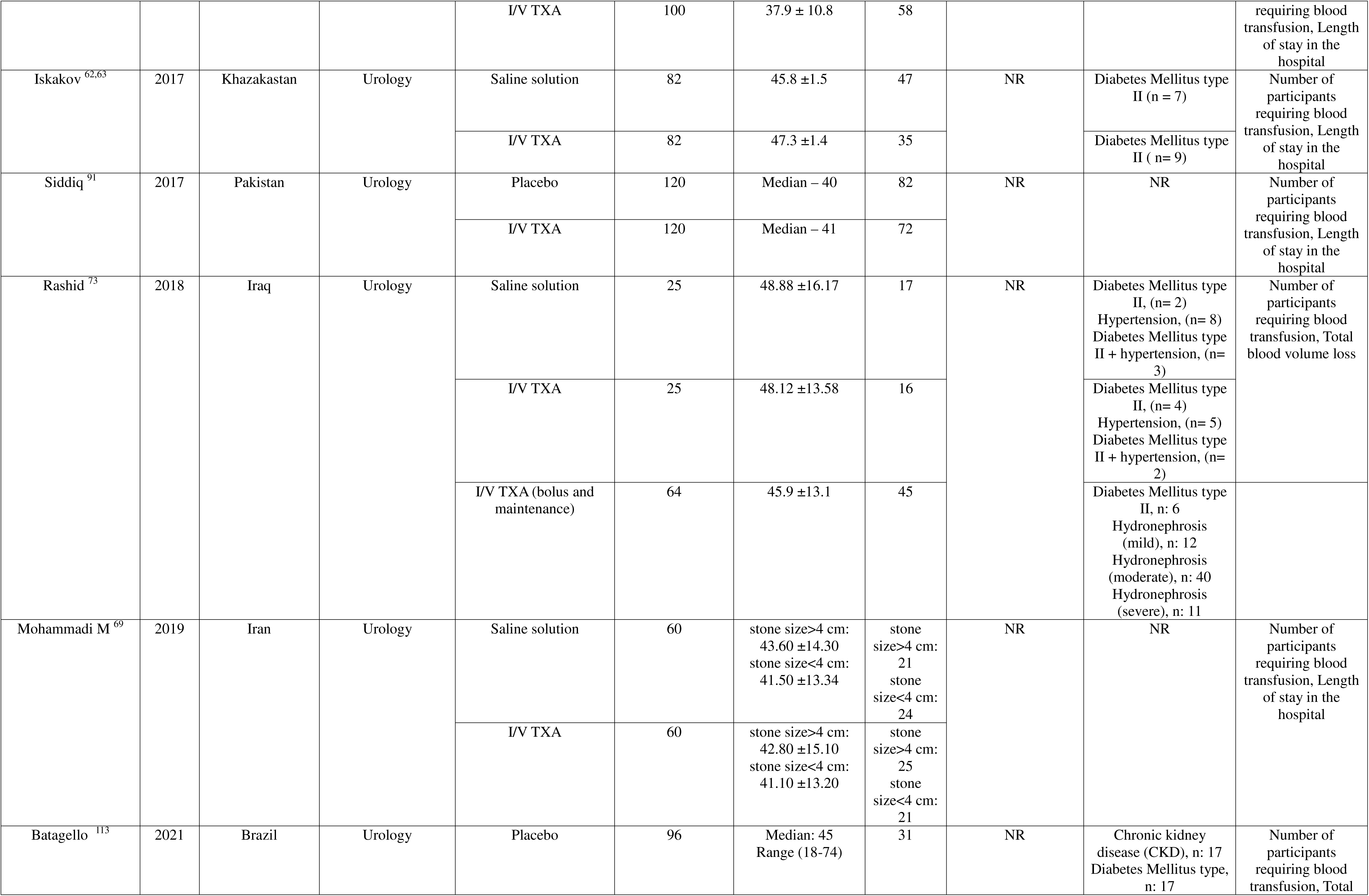
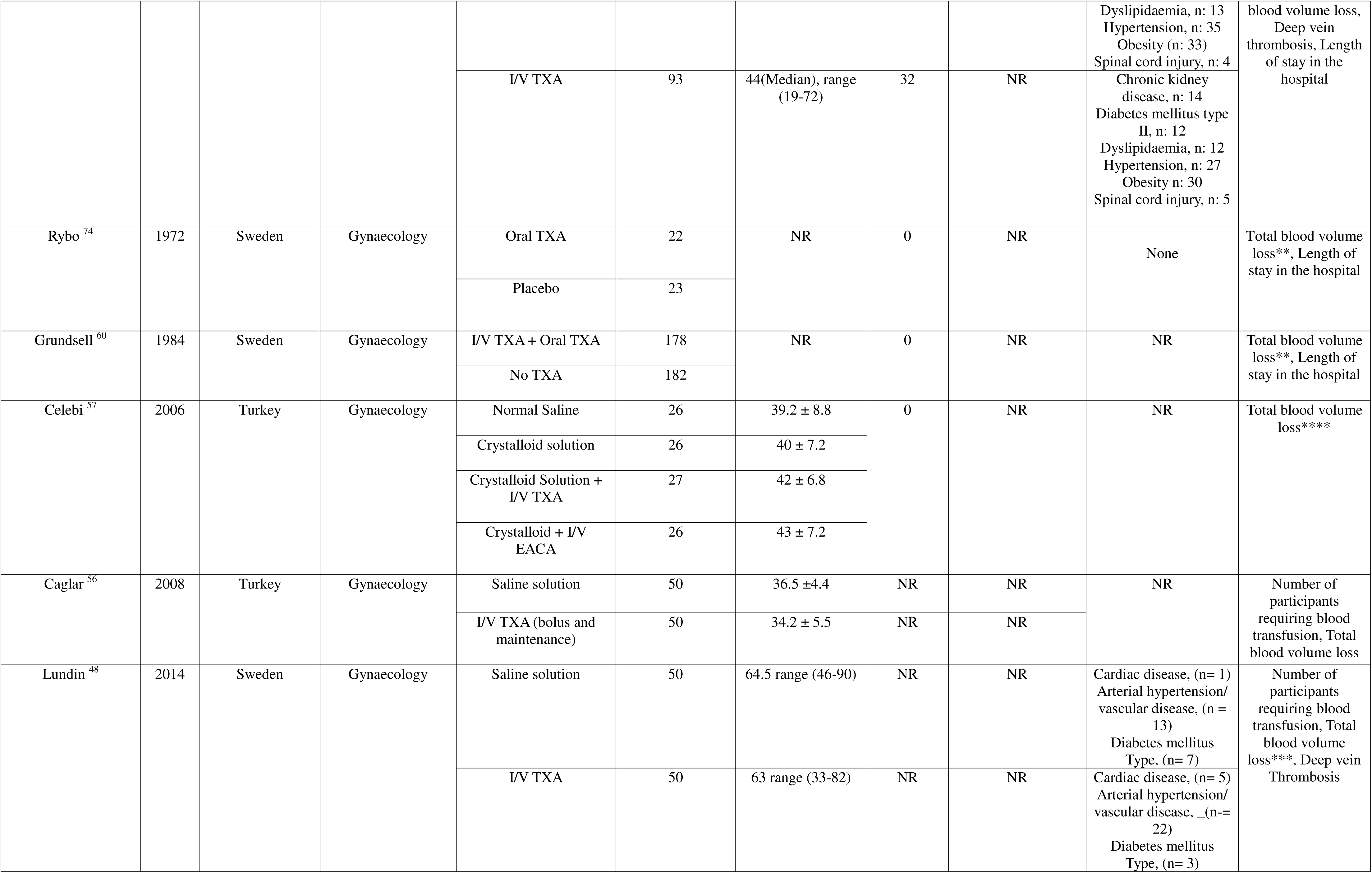
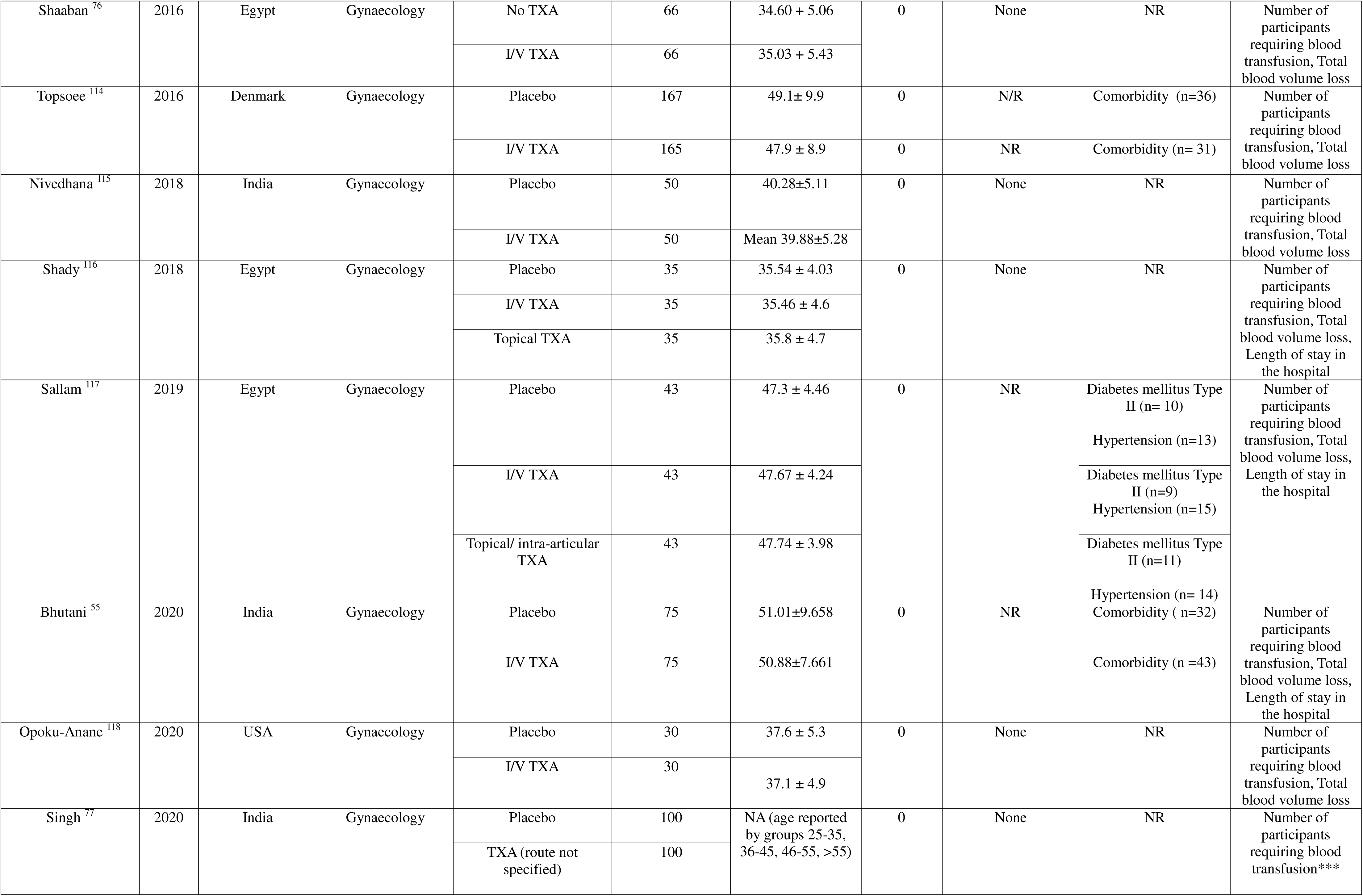
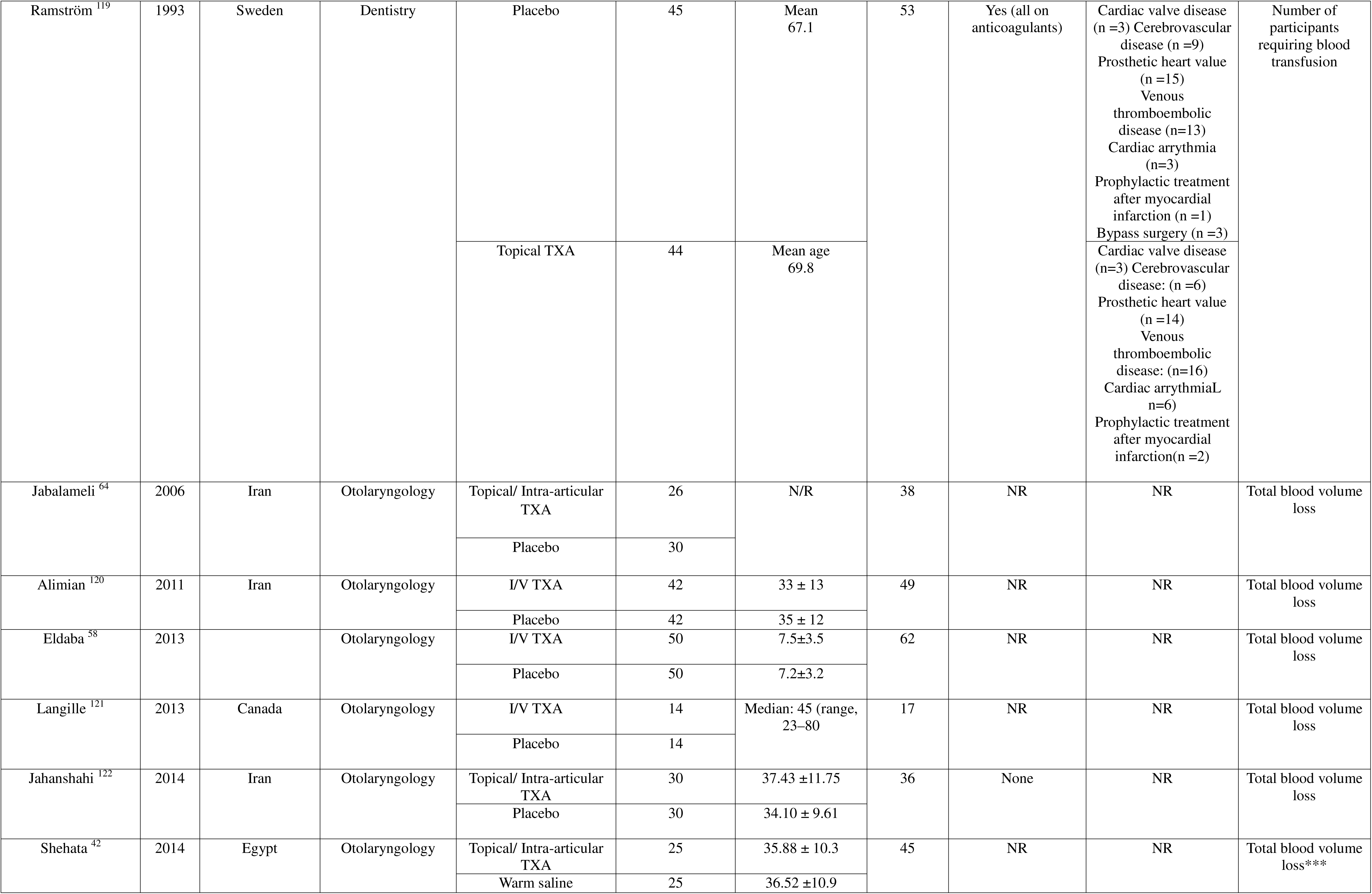
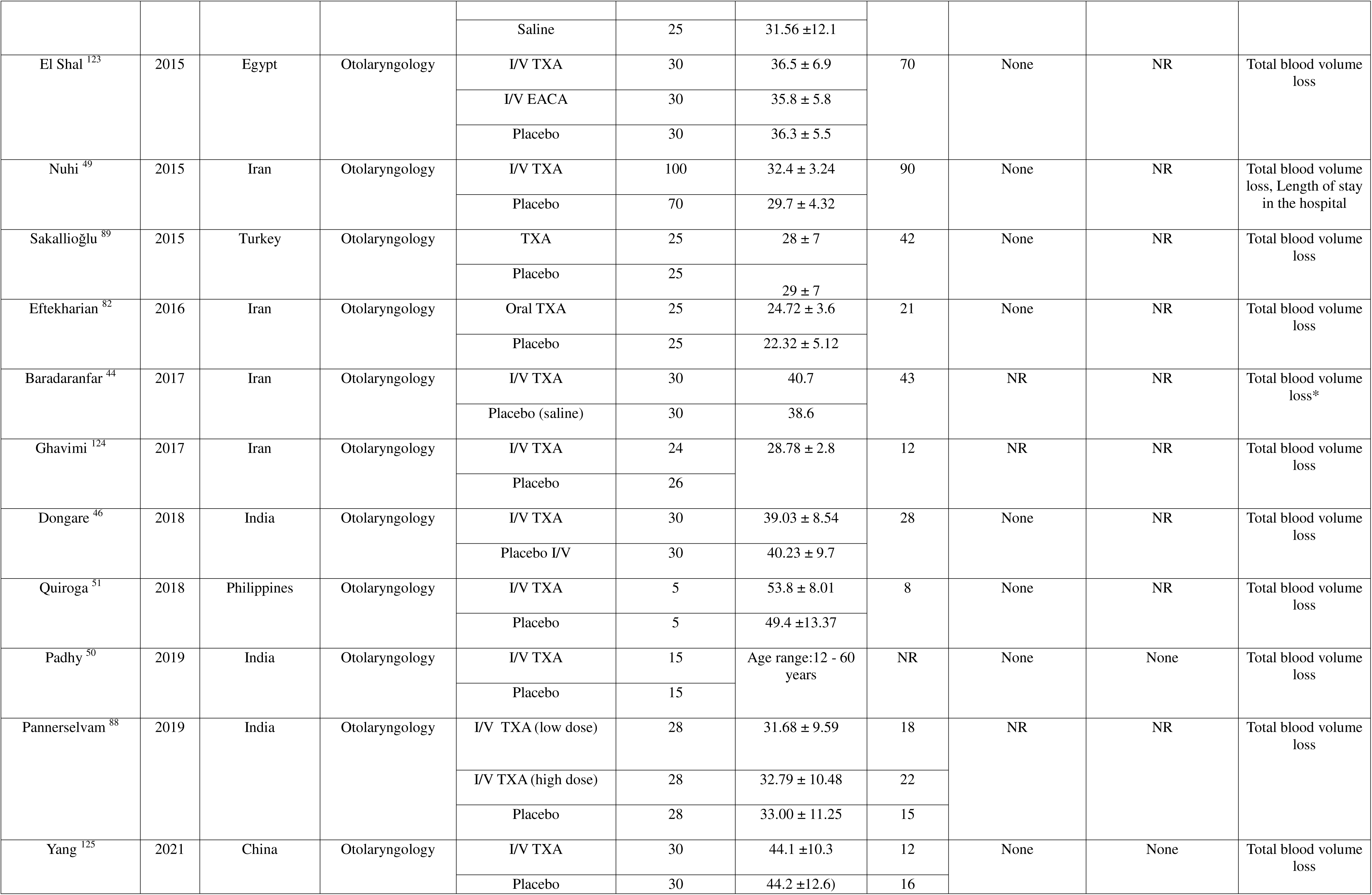
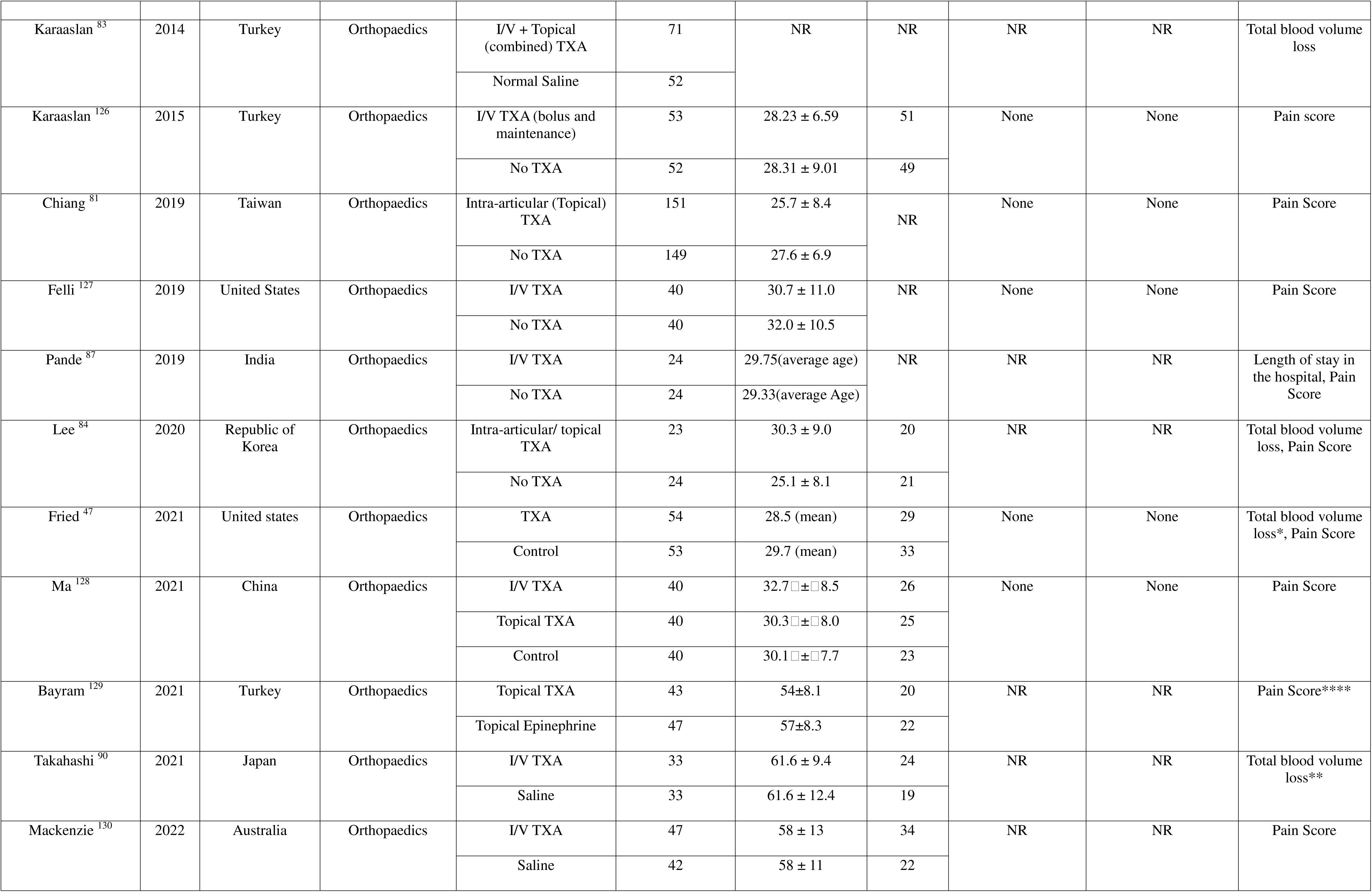
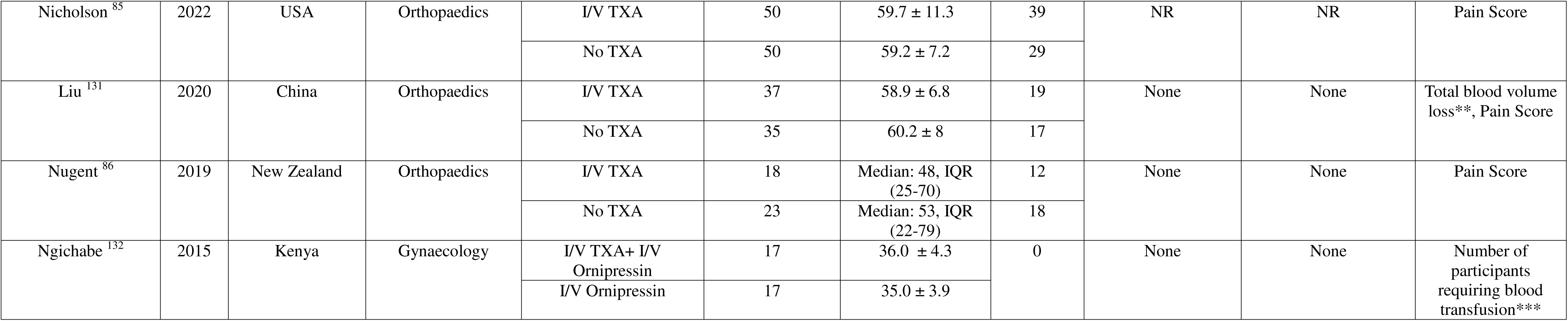
Characteristics of included studies (RCTs involving surgeries at low risk for blood loss) * = Outcome was excluded due to unobtainable standard deviations, ** = Outcome was excluded due to inability to access full text and thus data. *** = Outcome was excluded due to questioning validity of the reported data, **** = excluded due to no control arm.

There were 91 studies (n = 8887) identified from the orthopaedic specialty for any risk level of blood loss, comparing TBL in the TXA arm with control arm (Appendix 4, Table 1). These are considered in the section labelled explanatory analysis below.

### Meta-analysis for trials at low risk of blood loss

#### Primary Outcomes

##### Total Blood Loss (TBL)

There were 41 studies (n=4072) comparing total blood volume lost in the TXA arm with the control arm. Combining the trials in a pairwise random effects meta-analysis showed that the blood volume lost was less in the TXA arm compared to the control arm (RoM = 0·73, 95% CI 0·68, 0·79) suggesting 27% less blood loss in TXA arm (Figure 2A). There was large between-study variance (*tau* = 0·210; I^2^ = 93·69%). The funnel plots of the log RoM (appendix 3, fig 1) show many studies with similar standard errors closely clustered together, however, it is not perceived that there is a systematic exclusion of results from the non-significant region of the plot that would be indicative of publication bias. There is also no noticeable asymmetric pattern with respect to the labelled subgroups i.e. surgical speciality and route of TXA administration. GRADE assessment of the evidence base comparing the total blood loss during surgery in TXA and control groups was downgraded due to high between-study heterogeneity leading to low certainty of evidence (Table 3).

**Figure 2:**
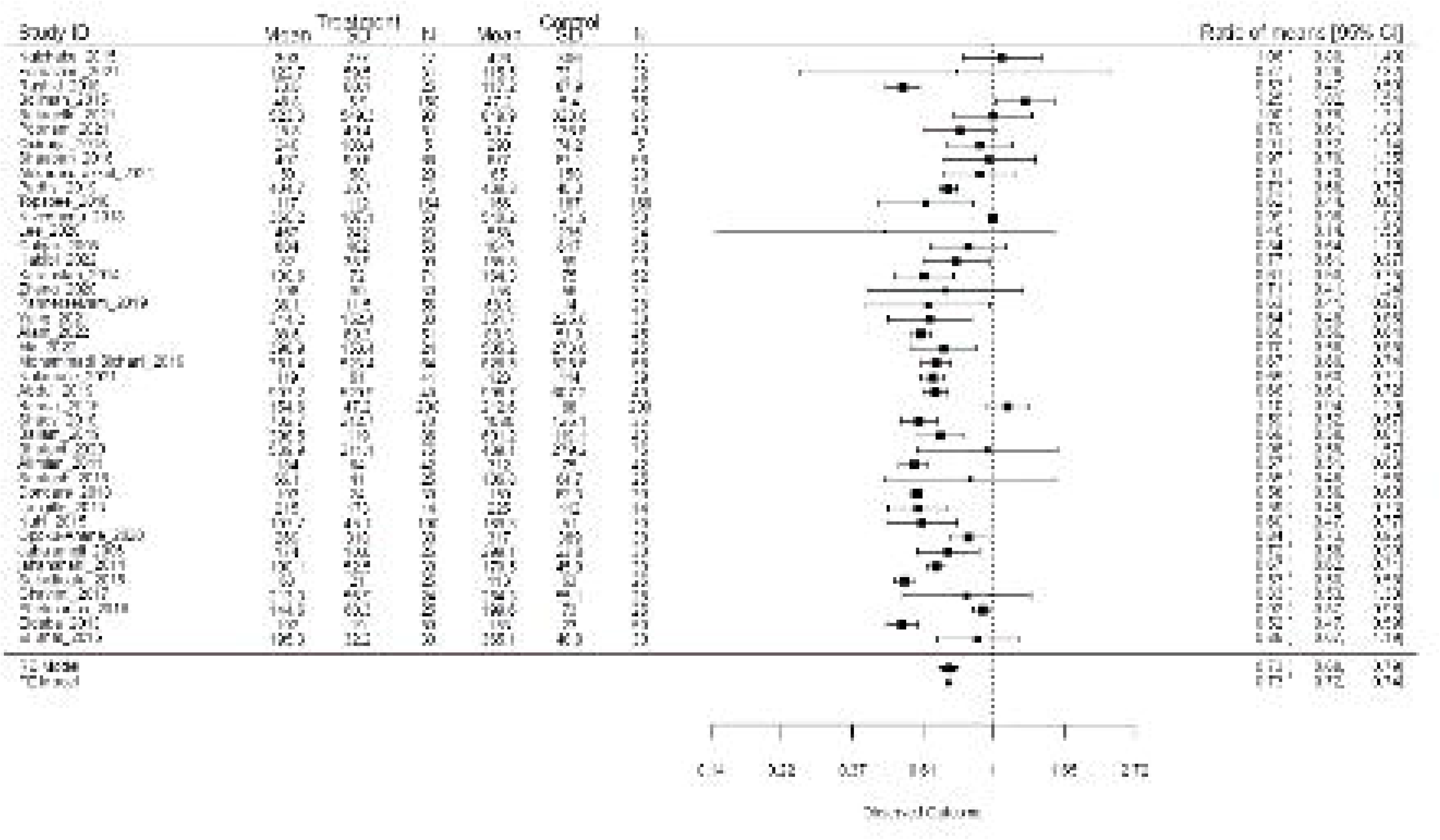
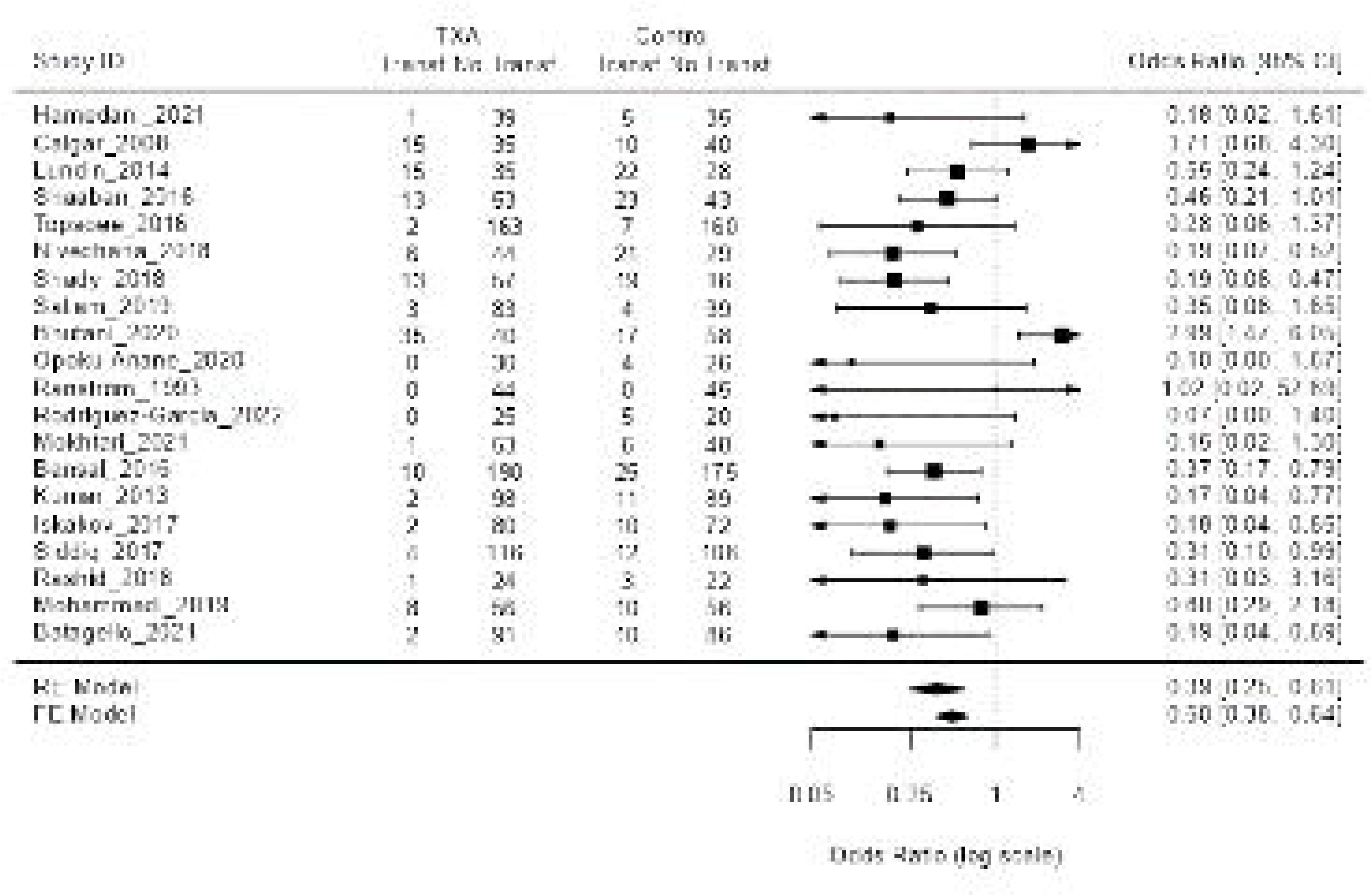
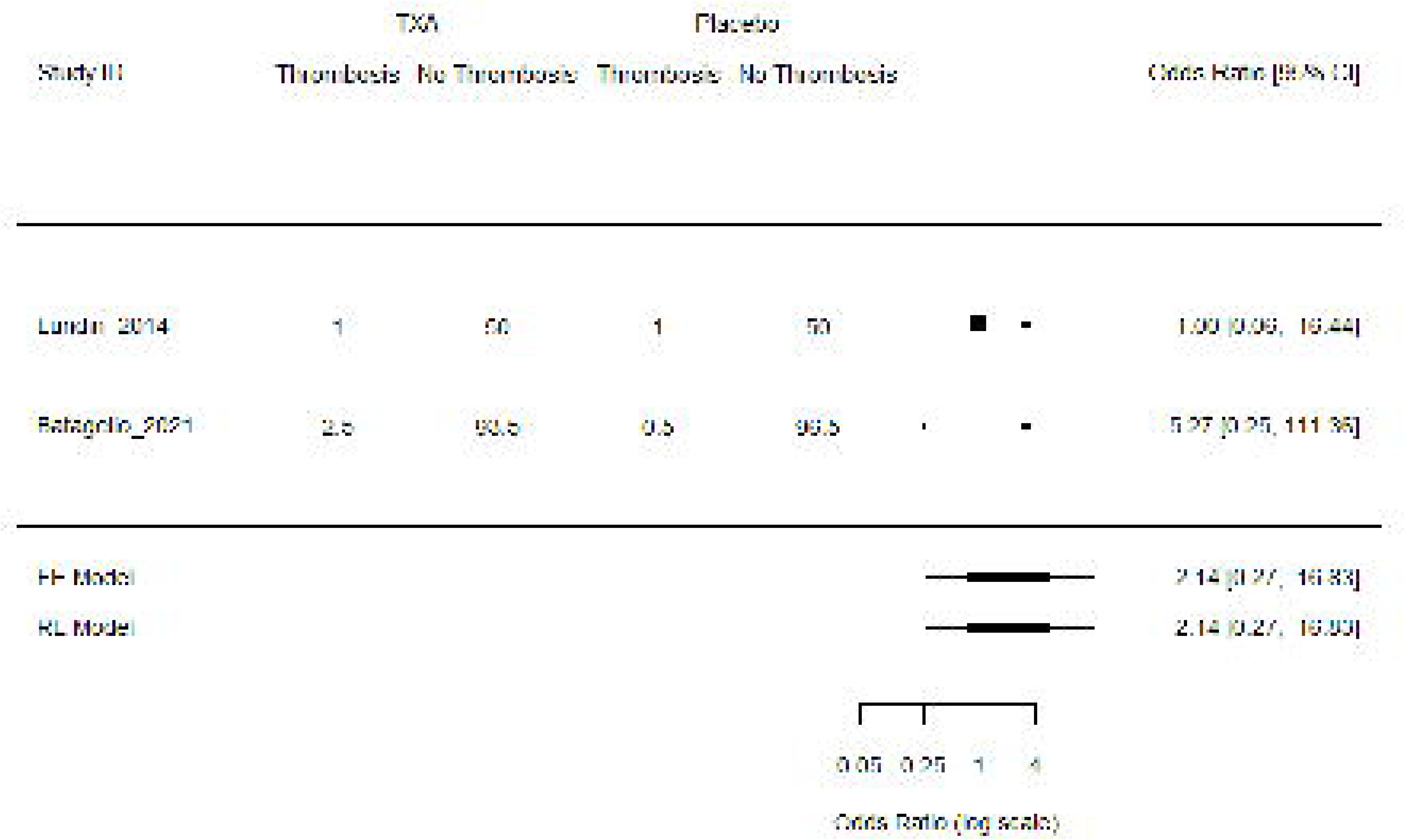
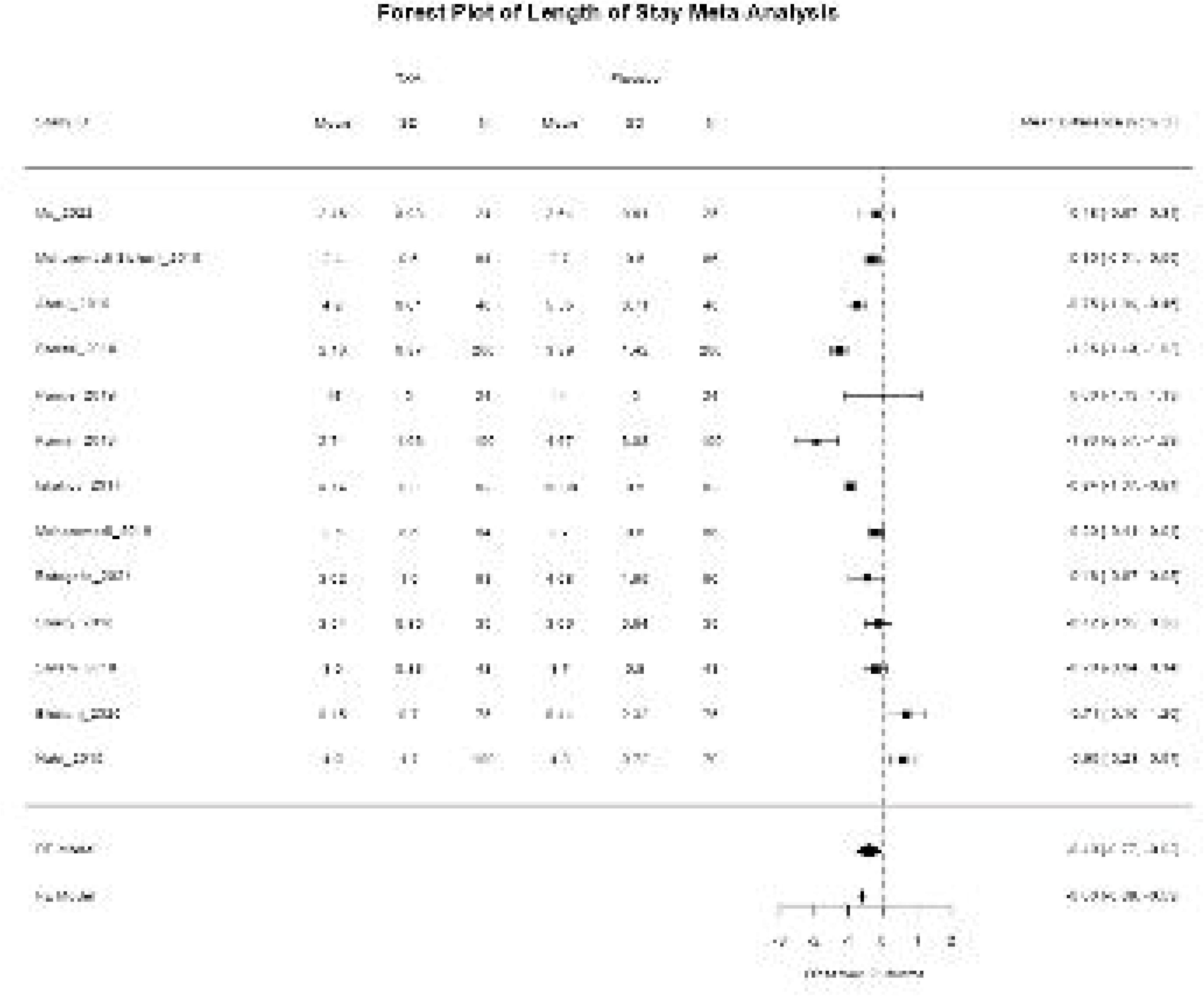
Forest plots showing pairwise meta-analysis for outcomes A- Total blood volume loss (ratio of means), B- Number of participants requiring blood transfusion (Odds ratio), CDeep vein thrombosis (Odds ratio), D- Length of stay in hospital

**Table 3:**
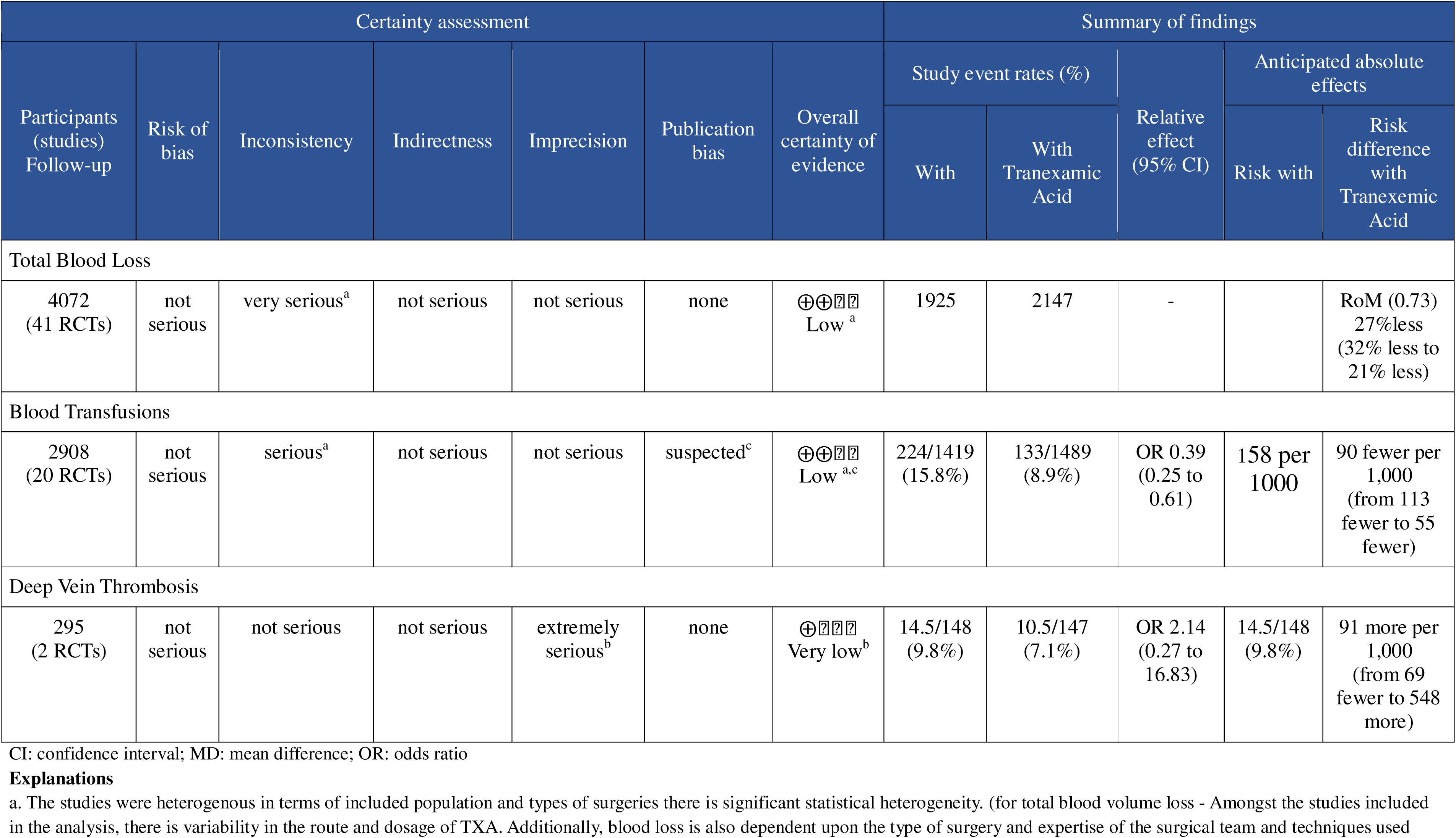
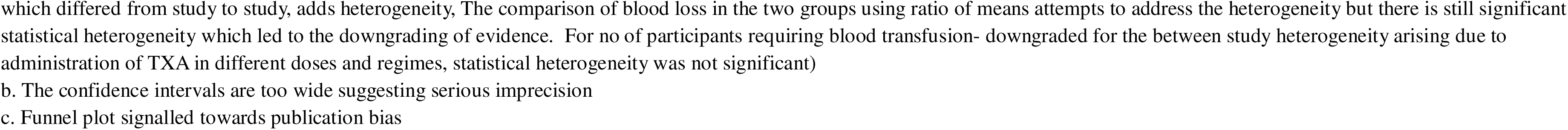
GRADE assessment and summary of findings table for primary outcomes.

| Certainty assessment |  |  |  |  |  |  | Summary of findings |  |  |  |  |
| --- | --- | --- | --- | --- | --- | --- | --- | --- | --- | --- | --- |
| Participants (studies)<br>Follow-up | Risk of bias | Inconsistency | Indirectness | Imprecision | Publication bias | Overall certainty of evidence | Study event rates (%) |  | Relative effect (95% CI) | Anticipated absolute effects |  |
|  |  |  |  |  |  |  | With | With Tranexamic Acid |  | Risk with | Risk difference with Tranexemic Acid |
| Total Blood Loss |  |  |  |  |  |  |  |  |  |  |  |
| 4072 (41 RCTs) | not serious | very serious<sup>a</sup> | not serious | not serious | none | ⊕⊕⊖⊖<br>Low <sup>a</sup> | 1925 | 2147 | - |  | RoM (0.73)<br>27% less<br>(32% less to 21% less) |
| Blood Transfusions |  |  |  |  |  |  |  |  |  |  |  |
| 2908 (20 RCTs) | not serious | serious<sup>a</sup> | not serious | not serious | suspected<sup>c</sup> | ⊕⊕⊖⊖<br>Low <sup>a,c</sup> | 224/1419 (15.8%) | 133/1489 (8.9%) | OR 0.39 (0.25 to 0.61) | 158 per 1000 | 90 fewer per 1,000 (from 113 fewer to 55 fewer) |
| Deep Vein Thrombosis |  |  |  |  |  |  |  |  |  |  |  |
| 295 (2 RCTs) | not serious | not serious | not serious | extremely serious<sup>b</sup> | none | ⊕⊖⊖⊖<br>Very low <sup>b</sup> | 14.5/148 (9.8%) | 10.5/147 (7.1%) | OR 2.14 (0.27 to 16.83) | 14.5/148 (9.8%) | 91 more per 1,000 (from 69 fewer to 548 more) |

CI: confidence interval; MD: mean difference; OR: odds ratio

##### Blood Transfusion

Of the 82 included studies, 20 (n= 2908) compared the number of participants requiring blood transfusions in the TXA arm with the control arm. The pairwise random effects meta-analysis indicated that the the odds of requiring blood transfusion was lower in the TXA arm compared to control arm (OR = 0·39, 95% CI = 0·25, 0·61) (Figure 2B). This suggests that the odds of requiring a transfusion is reduced by 61% with TXA. There was moderate between-study variance (*tau* = 0·754; I^2^ = 61·14%).

The funnel plots (Appendix 3, Figure 2) concluded that there was little evidence of publication bias. Using the GRADE approach, the certainty in the evidence for risk of blood transfusion was found to be low (Table 3).

The subgroup analyses (Appendix 3, Figure 3 & 4) for blood loss and transfusion outcomes found little indication that effectiveness of TXA varies by either type of surgery or route of administration.

**Figure 3:**
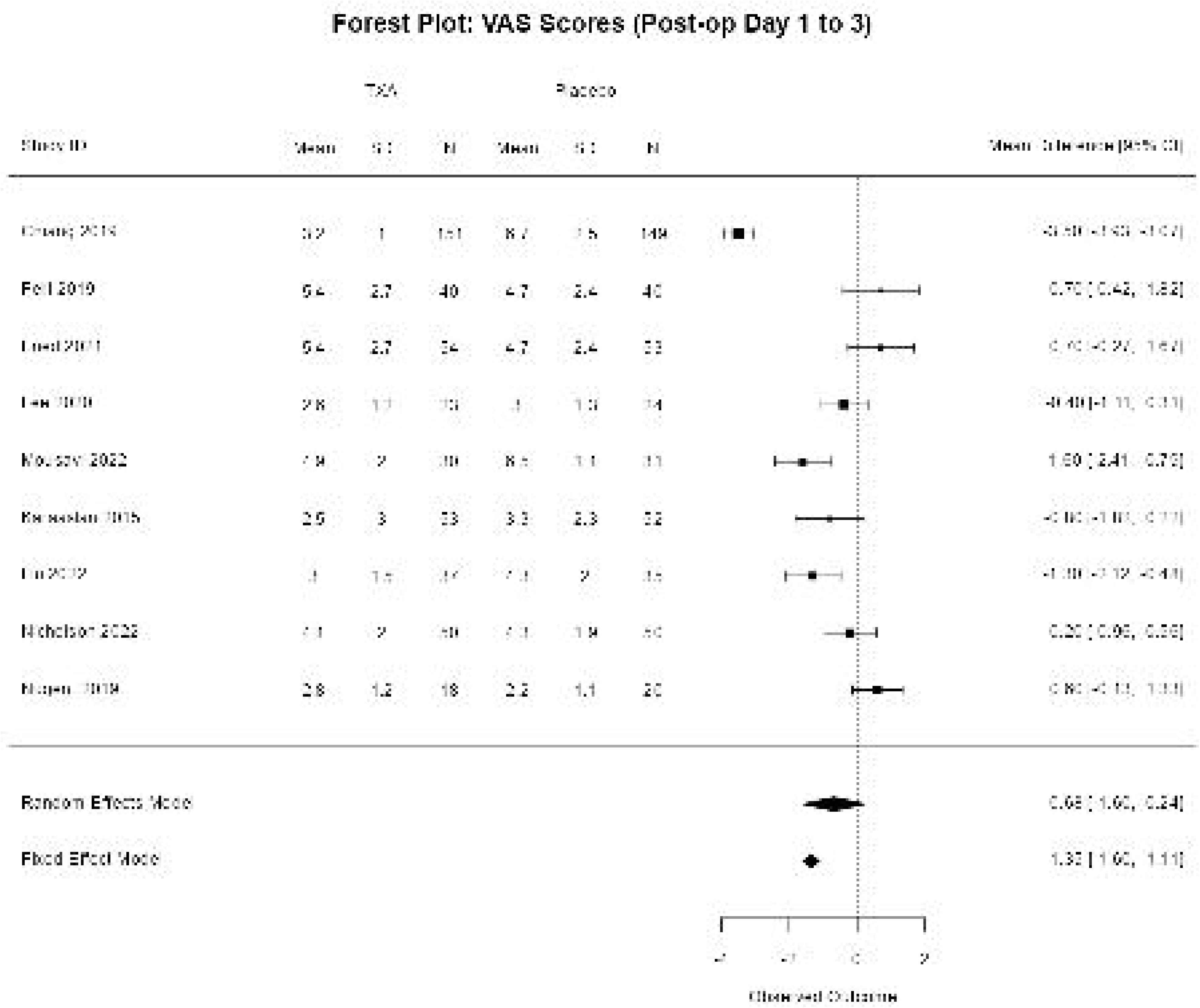
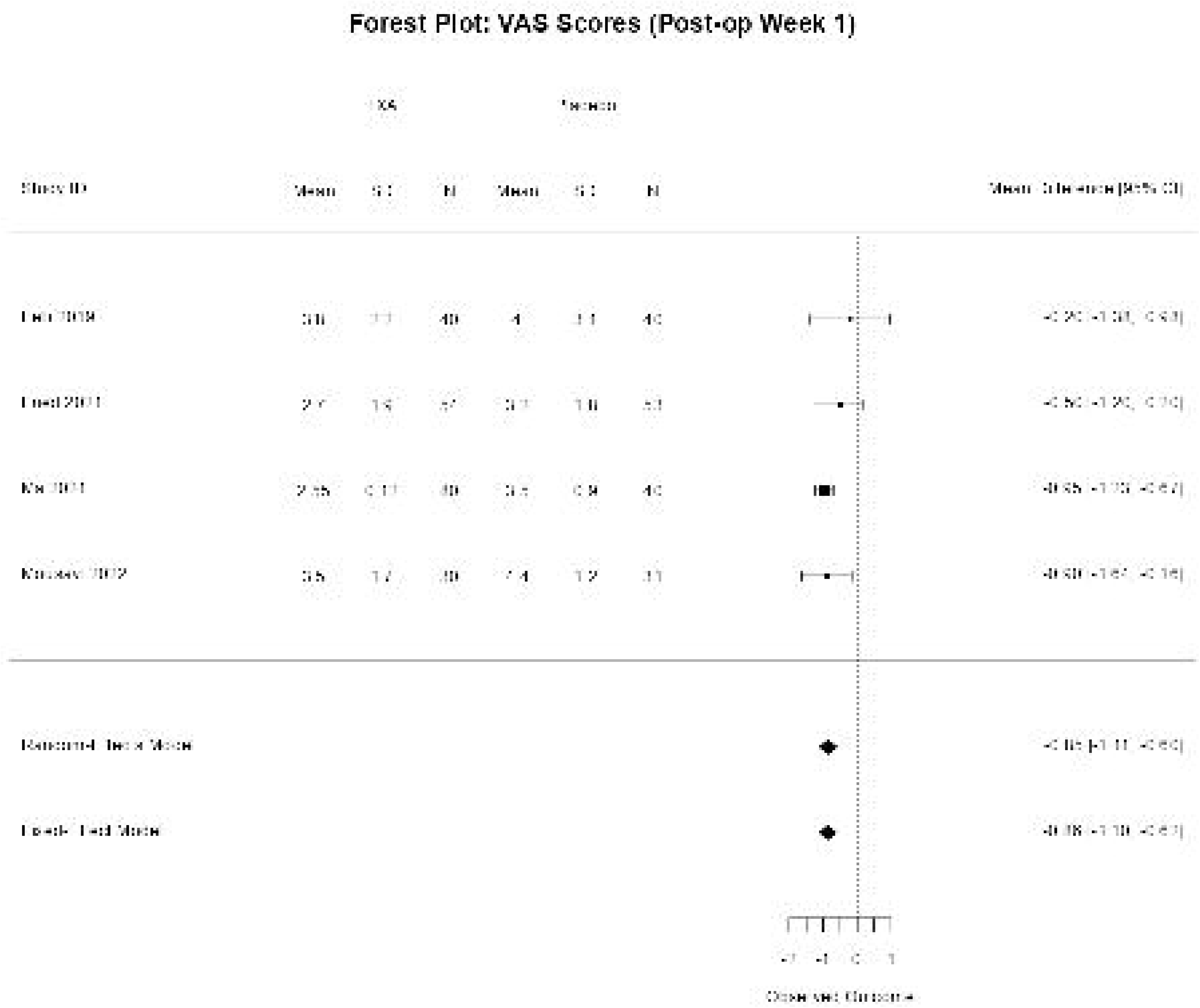
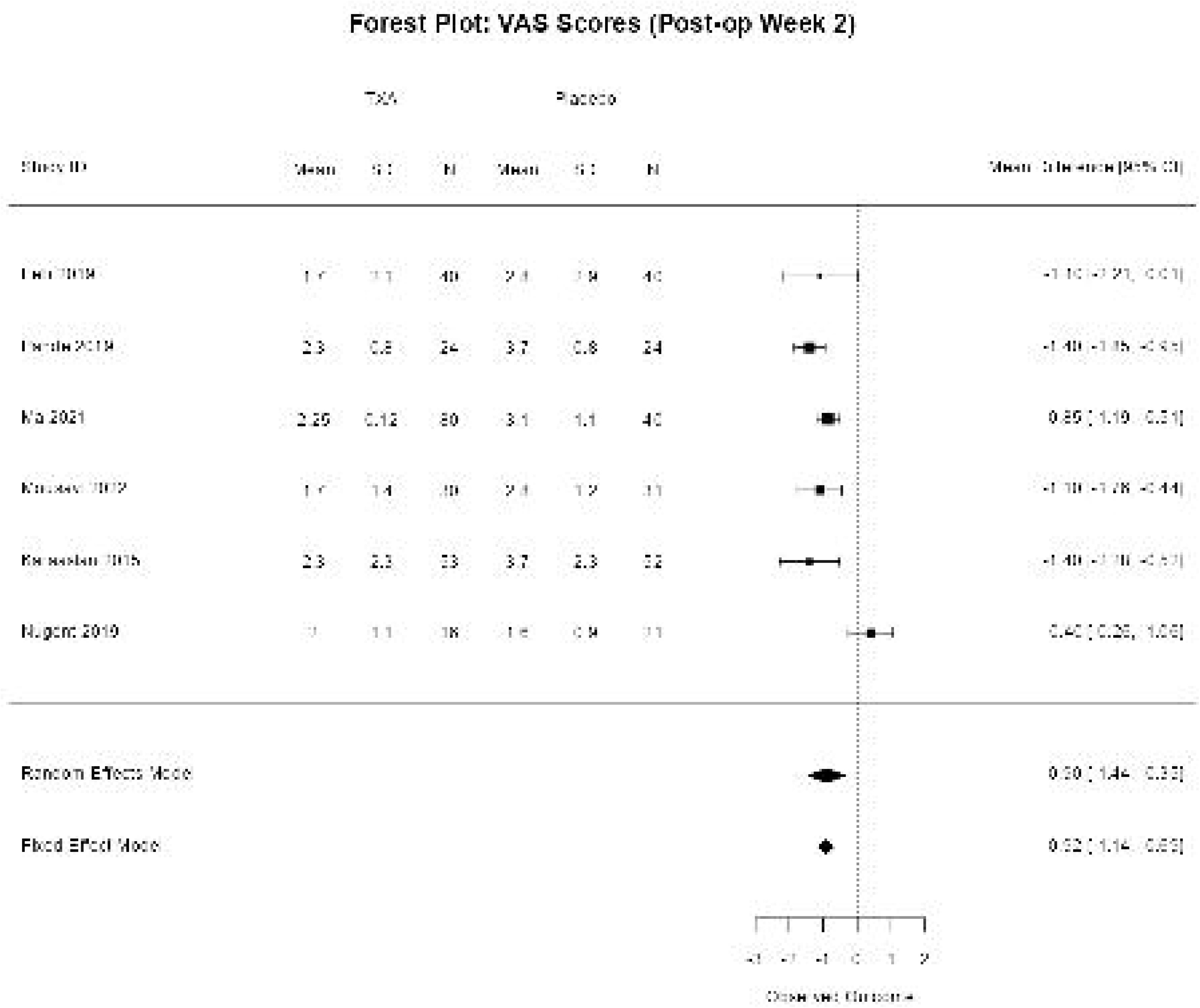
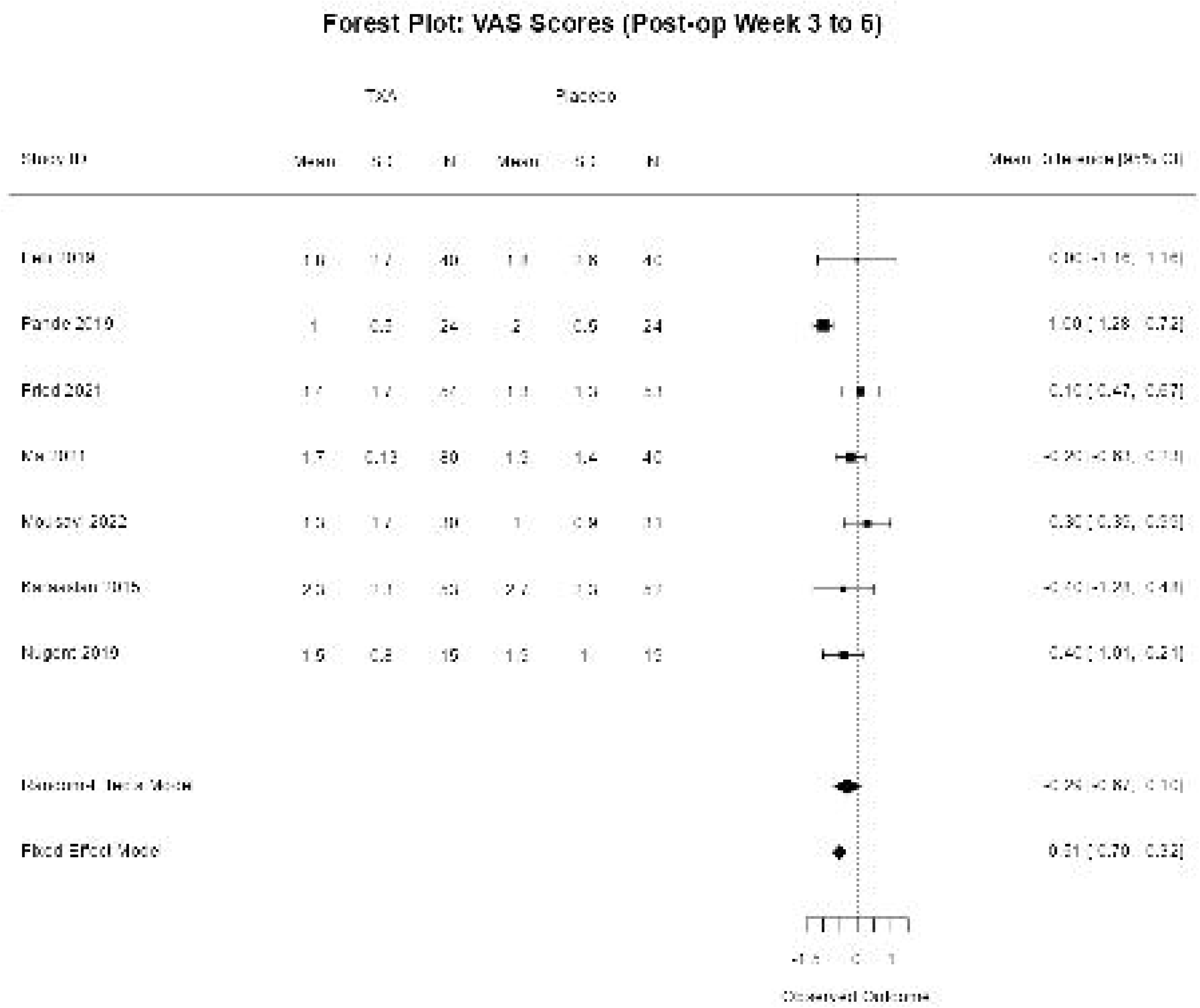
Forest plot for pairwise meta-analysis for outcome pain score at various timepoints A- post-op day 1-3, B- after post-op week 1, C- after post-op week 2, D- post-op week 3-6

##### Deep Vein Thrombosis (DVT)

Only two studies (n= 289) had data regarding any DVT events. Random effects meta-analysis for occurrence of DVT produced an odds ratio with a very wide confidence interval spanning no effect (OR 2·14, 95% CI 0·27, 16·83) (figure 2C). Furthermore, many trials stated that no thrombosis occurred in either study arm, and thus were not included in the quantitative analysis since trials with 0 total events do not contribute information on an odds ratio scale.

The GRADE assessment of the evidence base for DVT revealed very low certainty due to imprecision (Table 3).

#### Secondary outcomes

##### Length of Hospital Stay

There were 13 studies (n=1866) that reported length of hospital stay. The pairwise random effects meta-analysis estimated length of stay to be reduced by 0.4 days in the TXA group compared to control (MD = -0·40 days, 95% CI = -0·77, -0·02) (figure 2D).

##### Pain Score

There were 11 studies (n= 1041) that compared the visual analog scale (VAS) score for post-operative pain in the TXA arm with the control arm. The meta-analysis showed that initially (post-op day 1-3) there was no statistically significant difference in the mean pain score (MD = -0·68 95% CI -1·26, 0·24) (Figure 3A). However, by the end of post-operative week 1 TXA was associated with reduced pain (MD= -0·85, 95% CI -1·11, -0·60) (Figure 3B), which persisted at post-operative week 2 (MD = -0·90, 95% CI = -1·44, -0·35) (Figure 3C). Pain improvement in weeks 3 to 6, was smaller but still statistically significant (MD= -0·29, 95% CI –0·67, -0·10) (Figure 3D). The small and inconsistent number of studies contributing at each time point warrants caution when interpreting these findings.

##### Sensitivity Analyses

The results from the sensitivity analysis using fixed effect models were consistent with the results from the random effects models for all outcomes except length of hospital stay. Additonal sensitivity analysis for the length of hospital stay (Appendix 3, Figure 5) excluding a single influential trial^91^ (large effect size and small standard errors), found the random effects estimate remains largely unaffected and therefore conclusions do not change. However the magnitude of the fixed effect estimate is reduced dramatically from -0.60 (95% CI –0.68, -0.53) to –0.06 (-95% CI –0.09, -0.04).

### Exploratory Analysis (Including moderate-high risk trials from orthopaedics speciality) (Appendix 4)

#### Total Blood Loss

A meta-regression for mean difference of TBL, with TBL in the control arm as a covariate, had a regression slope gradient of -0·3 (95% CI -0·36, -0·23) (Appendix 4, figure 1). This suggests that for any increase in blood loss for a surgery TXA reduces the blood loss by approximately one third providing evidence that the difference in blood loss between groups is dependent on the blood loss in the control group. Adjusting the funnel plot using the residuals from a meta-analysis model that includes TBL in the control arm as a covariate substantially reduces the amount of asymmetry compared to the unadjusted funnel plot (Appendix 4, figure 2). The exploratory analysis also reinforces the decision to use RoM as a preferred outcome metric for the TBL outcome since it suggests a proportional, not absolute, difference in blood loss between groups across risk groups.

#### Blood Transfusion

Another meta-regression analysis examining the odds ratio for blood transfusion against control arm blood loss across low-risk (slope gradient= 0·0004 (p=0·68)), high-risk (slope gradient = -0·09 (p =0·40)), and combined risk groups (slope gradient = 0·03 (p=0·63)) suggested no statistically significant associations. This implies there is little evidence to suggest the treatment benefit of TXA varies systematically with the volume of blood loss (Appendix 4, figure 3). This analysis also highlights a large overlap in observed blood loss across trials - even within risk classifications – which probably reflects the challenges of predicting blood loss prior to an operation.

#### Interest-Holder Review

The authors met with clinical stakeholders on three occasions, complemented by email correspondence, and the public contributor group on two occasions. Both groups agreed that efforts to conserve blood resources were important. The clinical group felt that the body of evidence would support relaxing current limits on when to consider using TXA. They suggested that the current process of using thresholds of anticipated blood loss to trigger use of TXA may be overly complex and a barrier to use of TXA at scale. The public contributor group was keen that they would not be denied a treatment that may reduce adverse effects of surgery, although they expressed concerns about TXA if it were shown to increase thrombotic risk.

#### Cost Efectiveness Analysis

The clincal review found TXA reduced the transfusion rate in our low-risk studies from 17% to 7%, representing an average saving 0·095 units of blood per surgical procedure. TXA also reduced length of hospital stay by an average of 0·40 days, representing a cost-saving of £137 and a small QALY gain. Given modest TXA acqusition costs and reductions in blood transfusion, overall, TXA is modelled to save £156 per patient, with incremental net monetary benefit of £161 and 99% probability of being cost-effective at a £20,000 per QALY threshold (Appendix 5). Sensitivity and scenario analyses informed by the results of the clinical review had little impact on these results. Exclusion of any impact of length of stay had the greatest effect, though in this case TXA remained cost-effective even at very low levels of anticipated blood loss (Appendix 5).

## Discussion

Existing evidence and guidelines^5,6^ support the use of TXA in surgeries with an expected moderate or large blood loss. This review examines the clinical and cost-effectiveness of TXA in low-risk blood loss surgeries and found that TXA can significantly reduce blood loss and the odds of blood transfusion in surgeries classified as low-risk for blood loss. Although absolute blood loss and transfusion rates were low, the proportional benefits of TXA were consistent with findings from high-risk for blood loss surgeries, i.e. blood loss decreased by about one third and odds of blood transfusion decreased by 60% in the TXA group compared to control. Regarding safety, only two studies reported DVT, and pooled effects based on these studies suggested an increased risk but with large uncertainties. These findings do not provide a safety signal but point towards the need for ongoing pharmacovigilance and routine monitoring, should TXA use be expanded to low-risk surgeries. Additionally, TXA was associated with reduction in hospital stay and improvement in pain, thus offering further non-haemostatic benefits, possibly mediated through anti-inflammatory^92^ and tissue recovery effects.^93^

There is limited research on TXA in low-risk surgeries, but findings align with previous reviews of low-risk orthopedic surgeries^94^ and broader evidence on high-risk surgeries. ^95,96^ A key challenge in our work was defining the relevant evidence base for < 0.5 litre blood loss “low-risk” surgeries (since this is the current threshold recommended by NICE for use of TXA in the UK^5^). We based our inclusion criteria on a list of surgeries classified as low-risk using the ISTH guidance. However, for many included studies, the observed volumes of blood loss in the trial control arms were equal to, or greater than, surgeries classed as moderate- or high-risk of blood loss (i.e. >= 0·5 litres). Another challenge was addressing the proportional nature of the effect on blood volume loss, which has important implications for meta-analysis. We used methods to produce appropriate effect estimates, and this somewhat non-standard approach aligned with that of a previous meta-analysis.^95^

TXA has already been demonstrated cost-effective for surgeries where patients are at a moderate- or high-risk of bleeding. However, uncertainty in clinical assessment of bleed risk, a priori, and lack of analysis of the benefits of TXA for surgery with lower bleeding risk may contribute to underuse. Studies show that, even at the current threshold of 0·5 litres, a third of eligible UK patients do not receive TXA thus implying that as many as 33,000 transfusions could be prevented, saving 45,000 hospital days annually.^97,98^ Our analyses suggest TXA’s low cost and potential to shorten hospital stays make it cost-saving even for low-risk surgeries. Our analysis indicates TXA would remain cost-effective with transfusion risks as low as 2%.

The suboptimal implementation of guidance for use of TXA^99^ in moderate- to high-risk surgeries^99^ together with findings from this review, support expanding TXA use across all levels of bleeding risk. Broadening recommendations for TXA use would facilitate implementation of guidelines by removing the dependance on peri-operative prediction of blood loss.

### Strengths and limitations of the current review and economic analysis

A strength of this review is its comprehensive inclusion of evidence from all surgical specialties and various international healthcare systems, across all ages and without any limitations for comorbidities. Additionally, the review considered evidence in moderate to high risk of blood loss patients to better understand how the volume of blood lost without TXA impacted on clinical outcomes. Most studies were at low risk of bias, suggesting a robust evidence base. However, broad inclusion criteria introduced clinical heterogeneity. The analysis also hints towards potential publication bias in blood loss outcomes. The review does not address the optimal regime, dosage or route of administration for TXA, and confounders such as use of thromboprophylaxis or relevant morbidity were often not reported.

A substantial number of studies did not report primary outcomes, including blood loss and transfusion, which impilies that the trials used in each analysis may not represent the whole population. For surgical procedures involving otorhinolaryngology, dentistry, etc, blood transfusion is a rare event and is therefore seldom investigated. This restricted our analysis of transfusion to a limited number of surgical procedures.

Most of the studies did not find any thrombotic events, while this is suggestive of safety, zero event rates do not lend themselves to useful summary analyses. Secondary outcomes such as pain were not reported consistently and therefore limits the understanding of TXA’s broader clinical impact. A potential further limitation of our review was the reliance on previous systematic reviews for a proportion of the data extraction, where available, as we cannot verifythe quality of the extracted data. Although the economic analysis focussed on short-term hospital costs, any post-discharge benefits would only strengthen TXA’s economic case. Adverse effect of TXA, such as thrombosis, would be expected to carry longer term consequences. However, we did not include any such effects based on the evidence from the clinical review.

### Implications for Research and Practice

The review recommends that future studies of TXA prioritise non-bleeding outcomes, particularly in surgeries with a low-risk of bleeding, to better explore and understand its broader clinical impact. Additionally, future research on TXA use in this group of surgeries, should evaluate optimal dosing regimens and routes of administration.

The findings from the review suggest TXA is clinically effective and cost-effective, consistently reducing bleeding-related outcomes and other patient-reported outcomes. Our results indicate that current TXA guidelines should be reviewed to include surgeries with any potential for blood loss.

## Data Availability

Datasets are available on reasonable request to the corresponding author.

## Acknowledgements

We are grateful to the Evidence Synthesis Public Involvement Group (ESPIG) who collarobated on this project for their thoughtful contributions, which informed the development and focus of this review. We also wish to thank the clinicians we contacted who provided expert input and advice throughout the review process. We are grateful to colleagues from the NHS Blood and Transfusion Systematic Review Initiative for their advice which helped shaped the review.

## Role of Funding Agency

This study is funded by the NIHR [NIHR Evidence Synthesis programme (NIHR153934)]. The views expressed are those of the authors’ and not necessarily those of the NIHR or the Department of Health and Social Care.

## Authorship Contributions

AS-Conceptualisation, Methodology, Supervision, Writing-review and editing

NC-Conceptualisation, Methodology, Supervision, Writing-review and editing

AD-Conceptualisation, Methodology, Supervision, Writing-review and editing

OW-Conceptualisation, Methodology, Supervision, Writing-review and editing

TQ-Conceptualisation, Methodology, Supervision, Writing-review and editing

ANS-Literature Search, Methodology, Writing-review and editing

NJ-Data Collection, Data Interpretation, Writing-original draft

WR-Data Collection, Formal Analysis, Writing-review and editing

GC-Data Collection, Formal Analysis, Writing-review and editing

HT-Data Collection, Formal Analysis

ML-Data Collection

RM-Data Collection, Writing-review and editing

CN-Data Collection, Writing-review and editing

TM-Data Collection, Formal Analysis

EF-Data Collection, Formal Analysis, Writing-review and editing

