## Supplementary file for "Tranexamic acid in reducing expected blood loss in moderate to low risk surgeries: systematic review, meta-analysis and cost effectiveness analysis"

**Appendices**

**Appendix 1**

**Table 1: Detailed Search Strategy for databases- Cochrane CENTRAL, Medline , Embase, CDSR- Cochrane database of systematic reviews, clinicaltrials.gov and WHO ICTRP**

| **Source** | **Search strategy** |
| --- | --- |
| 1. CENTRAL (The Cochrane Library via CRS)       Most recent search: 21/06/2023 | #1 tranexamic 3852  #2 MeSH descriptor: [Tranexamic Acid] explode all trees 1525  #3 TXA 1285  #4 tranexmic 2  #5 tranhexamic 9  #6 #1 or #2 or #3 or #4 or #5 3905  #7 surgery 296524  #8 surgical 130415  #9 operation 49097  #10 operations 9798  #11 surgeries 13364  #12 operative 55355  #13 post-operative 26001  #14 postoperative 148162  #15 pre-operative 6535  #16 preoperative 45623  #17 hospitalised 21673  #18 hospitalized 21673  #19 MeSH descriptor: [General Surgery] explode all trees 501  #20 #7 or #8 or #9 or #10 or #11 or #12 or #13 or #14 or #15 or #16 or #17 or #18 or #19 395300  #21 #20 and #6 |
| 2. MEDLINE (Ovid SP)  1946 – 20 June 2023    Most recent search: 21/06/2023 | 1. tranexamic.ti,ab,kw.  2. exp Tranexamic Acid/  3. TXA.ti,ab,kw.  4. Tranhexamic.ti,ab,kw.  5. tranexmic.ti,ab,kw.  6. or/1-5  7. surgery.ti,ab,kw.  8. surgical.ti,ab,kw.  9. surgeries.ti,ab,kw.  10. operation.ti,ab,kw.  11. operations.ti,ab,kw.  12. post-operative*.ti,ab,kw.  13. pre-operative*.ti,ab,kw.  14. procedure*.ti,ab.  15. theatre.ti,ab.  16. exp General Surgery/  17. Postoperative Complications/  18. postoperative.ti,ab,kw.  19. preoperative.ti,ab,kw.  20. or/7-19  21. 6 and 20  22. randomized controlled trial.pt.  23. controlled clinical trial.pt.  24. randomized.ab.  25. placebo.ab.  26. clinical trials as topic.sh.  27. randomly.ab.  28. trial.ti.  29. or/22-28  30. exp animals/ not humans.sh.  31. 29 not 30  32. 21 and 31  33. (2015* or 2016* or 2017* or 2018* or 2019* or 2020* or 2021* or 2022* or 2023*).ed.  34. (2015* or 2016* or 2017* or 2018* or 2019* or 2020* or 2021* or 2022* or 2023*).yr.  35. 33 or 34  36. 32 and 35 |
| 3. Embase (Ovid SP)  1996 to 2023 Week 24    Most recent search: 21/06/2023 | 1. tranexamic.ti,ab,kw.  2. exp tranexamic acid/  3. TXA.ti,ab,kw.  4. Tranhexamic.ti,ab,kw.  5. tranexmic.ti,ab,kw.  6. or/1-5  7. surgery.ti,ab,kw.  8. surgical.ti,ab,kw.  9. surgeries.ti,ab,kw.  10. operation.ti,ab,kw.  11. operations.ti,ab,kw.  12. post-operative*.ti,ab,kw.  13. pre-operative*.ti,ab,kw.  14. procedure*.ti,ab.  15. theatre.ti,ab.  16. exp elective surgery/ or exp general surgery/ or exp surgery/  17. postoperative care/ or postoperative complication/ or postoperative period/ or postoperative hemorrhage/  18. postoperative.ti,ab,kw.  19. preoperative.ti,ab,kw.  20. or/7-19  21. 6 and 20  22. randomized controlled trial/  23. controlled clinical trial/  24. randomized.ab.  25. placebo.ab.  26. randomly.ab.  27. trial.ti.  28. randomised.ab.  29. double-blind*.ti,ab.  30. single-blind*.ti,ab.  31. or/22-30  32. 21 and 31  33. (2015* or 2016* or 2017* or 2018* or 2019* or 2020* or 2021* or 2022* or 2023*).dc.  34. (2015* or 2016* or 2017* or 2018* or 2019* or 2020* or 2021* or 2022* or 2023*).yr.  35. 33 or 34  36. 32 and 35 |
| 4. CDSR    Most recent search: 21/06/2023 | #1 tranexamic 3852  #2 MeSH descriptor: [Tranexamic Acid] explode all trees 1525  #3 TXA 1285  #4 tranexmic 2  #5 tranhexamic 9  #6 #1 or #2 or #3 or #4 or #5 3905  #7 surgery 296524  #8 surgical 130415  #9 operation 49097  #10 operations 9798  #11 surgeries 13364  #12 operative 55355  #13 post-operative 26001  #14 postoperative 148162  #15 pre-operative 6535  #16 preoperative 45623  #17 hospitalised 21673  #18 hospitalized 21673  #19 MeSH descriptor: [General Surgery] explode all trees 501  #20 #7 or #8 or #9 or #10 or #11 or #12 or #13 or #14 or #15 or #16 or #17 or #18 or #19 395300  #21 #20 and #6 |
| 5. ClinicalTrials.gov    Most recent search: 06/06/2023 | Advanced search:  (Tranexamic OR TXA) |
| 6. ICTRP (WHO Search Portal)    Most recent search: 06/06/2023 | Advanced search:  (Tranexamic OR TXA) AND surgery |

**Appendix 2**

**Risk of bias assessment**

**Table- 1 Criteria used for risk of bias assessment**

| **Domain** | **Rating** | **Criteria** |
| --- | --- | --- |
| **Random sequence generation** | Low risk of bias | Study used a random method, such as a computer‐generated system or random number table, to generate the allocation sequence and described the approach in sufficient detail. Drawing of lots, tossing of coin, shuffling of cards, or throwing dice was considered adequate if a person who was not otherwise involved in the recruitment of participants performed the procedure. |
|  | Unclear risk of bias | Methods used to generate the allocation sequence were not described in sufficient details. |
|  | High risk of bias | Study used a non‐random method, such as dates, names, or admittance numbers, to generate the allocation sequence of participants. |
| **Allocation concealment** | Low risk of bias | Allocation of participants to study groups was concealed from participants and investigators using a central independent unit, on‐site locked computer, identical syringes or schedules (used by an independent pharmacist or investigator), or opaque, sealed envelopes. |
|  | Unclear risk of bias | Methods for allocation concealment were not described or sufficient details were not provided to permit a judgement of low or high risk of bias. |
|  | High risk of bias | Allocations were not concealed and were known to either participants and investigators or both. |
| **Blinding of participants and personnel** | Low risk of bias | Participants and investigators were blinded; blinding was described in sufficient detail; and it was unlikely that the blinding could have been broken. |
|  | Unclear risk of bias | Blinding was not described or was not described in sufficient detail to permit a judgement of low or high risk of bias. |
|  | High risk of bias | Participants or investigators were not blinded; blinding was broken; or it was likely that the outcome could have been affected by the lack of blinding. |
| **Blinding of outcome assessment** | Low risk of bias | Outcome assessment was blinded; details of blinding were described in sufficient detail; and it was unlikely that the blinding could have been broken, or there was no blinding, but the outcome assessment was unlikely to have been affected by the lack of blinding. |
|  | Unclear risk of bias | Blinding was not described or was not described in sufficient detail to permit a judgement of low or high risk of bias. |
|  | High risk of bias | Outcome assessment was not blinded; blinding was broken; or it was likely that the outcome assessment could have been influenced by the lack of blinding. |
| **Incomplete outcome data** | Low risk of bias | No missing data: the reasons for the missing data were unrelated to the true outcome; or the study used appropriate methods to impute the data |
|  | Unclear risk of bias | Insufficient information to permit a judgement of low or high risk of bias |
|  | High risk of bias | Reasons for missing data were related to the true outcome, or the study used inappropriate methods to impute the data. |
| **Selective outcome reporting** | Low risk of bias | Study protocol was available, and all prespecified outcomes were reported and, in the manner, specified; or study protocol was not available, but it was clear that all prespecified outcomes had been reported. |
|  | Unclear risk of bias | Insufficient information to permit a judgement of low or high risk of bias |
|  | High risk of bias | Study protocol was available, but not all of the study’s prespecified outcomes were reported, or not all were reported in prespecified way, or one or more were reported incorrectly; or outcomes were reported that were not prespecified; or study did not have a protocol, and not all expected outcomes were reported. |

**Table 2—Results of risk of bias assessment for the included studies**

| **Study** | **Random sequence generation** | **Allocation concealment** | **Blinding of participants and personnel** | **Blinding of outcome assessment** | **Incomplete outcome data** | **Selective reporting** |
| --- | --- | --- | --- | --- | --- | --- |
| Hamedani 2021^59^ | Unclear | Unclear | Unclear | Unclear | Unclear | Low |
| Akkaranurakkul 2021 ^101^ | Low | Low | Low | Low | Low | Low |
| Mitra 2022 ^68^ | Unclear | Unclear | Unclear | Unclear | Low | Low |
| Hazrati 2021 ^61^ | Low | Unclear | Low | Low | Low | Low |
| Fornazieri 2021 ^100^ | Low | Low | Low | Low | Low | Low |
| Farsani 2022 ^102^ | Low | Low | Low | Low | Low | Low |
| Abtahi 2023 ^103^ | Low | Low | Low | Low | Low | Low |
| Habibi 2022 ^104^ | Low | Low | Low | Low | Low | Low |
| Volodymyr 2021 ^79^ | Unclear | Unclear | Unclear | Unclear | Unclear | Low |
| Salamah 2023 ^105^ | Low | Low | Low | Low | Low | Low |
| Shafa 2022 ^106^ | Low | Low | Low | Low | Low | Low |
| Alam 2022 ^54^ | Low | Low | Low | Unclear | Low | Low |
| Nalamate 2022 ^72^ | Low | Low | Low | Low | Low | Unclear |
| Abdul 2019 ^109^ | Low | Low | Low | Low | Low | Low |
| Aboelsuod 2023 ^110^ | Low | Low | Low | Low | Low | Low |
| Ahmadi 2023 ^43^ | Low | Low | Low | High | Low | Low |
| Abianeh 2022 ^53^ | Low | Low | Low | Unclear | Low | Unclear |
| Soliman 2015 ^78^ | Unclear | Unclear | Unclear | Unclear | Low | Unclear |
| Santosh 2016 ^75^ | Unclear | Unclear | Unclear | Unclear | Low | Unclear |
| Kulkarni 2019 ^65^ | Low | Unclear | Low | Low | Low | Unclear |
| Hamed 2020 ^107^ | Low | Low | Low | Low | Low | Low |
| Rodriguez-Garcia 2022 ^52^ | Low | Low | High | High | Low | Low |
| Ma 2022 ^67^ | Unclear | Unclear | Low | Low | Low | Low |
| Zhang 2020 ^80^ | Low | Unclear | Low | Low | Low | Low |
| Choudhury 2021 ^45^ | High | High | Unclear | Unclear | Low | Low |
| Mohammadi Sichani 2019 ^108^ | Low | Low | Low | Low | Low | Low |
| Mokhtari 2021^70^ | Unclear | Unclear | Low | Low | Low | Low |
| Bansal 2017 ^111^ | Low | Low | Low | Low | Low | Low |
| Mousavi 2022 ^71^ | Unclear | Unclear | Low | Low | Low | Low |
| Poonam 2021 ^112^ | Low | Low | Low | Low | Low | Low |
| Kumar, 2013 ^66^ | Low | Low | Unclear | Unclear | Low | Low |
| Iskakov, 2017 ^62,63^ | Low | Unclear | Unclear | Low | Unclear | Low |
| Siddiq, 2017 ^91^ | Low | Low | Low | Low | Low | Low |
| Rashid, 2018 ^73^ | Low | Low | Low | Low | Unclear | Unclear |
| Mohammadi M, 2019 ^69^ | Low | Low | Low | Unclear | Low | Low |
| Batagello, 2021 ^113^ | Low | Low | Low | Low | Low | Low |
| Rybo, 1972 ^74^ | Unclear | Unclear | Unclear | Unclear | Low | Unclear |
| Grundsell, 1984 ^60^ | Unclear | Unclear | Unclear | Unclear | Low | Unclear |
| Celebi, 2006 ^57^ | Unclear | Unclear | Unclear | Unclear | Low | Low |
| Caglar, 2008 ^56^ | Low | Low | Low | Low | Unclear | Low |
| Lundin, 2014 ^48^ | Low | Low | Low | Low | High | Low |
| Ngichabe, 2015 ^132^ | Low | Low | Low | Low | Low | Low |
| Shaaban, 2016 ^76^ | Low | Low | Unclear | Unclear | Low | Low |
| Topsoee, 2016 ^114^ | Low | Low | Low | Low | Low | Low |
| Nivedhana, 2018 ^115^ | Low | Low | Low | Low | Low | Low |
| Shady, 2018 ^116^ | Low | Low | Low | Low | Low | Low |
| Sallam, 2019 ^117^ | Low | Low | Low | Low | Low | Low |
| Bhutani, 2020 ^55^ | Low | Unclear | Unclear | Low | Low | Low |
| Opoku-Anane, 2020 ^118^ | Low | Low | Low | Low | Low | Low |
| Singh, 2020 ^77^ | Unclear | Unclear | Unclear | Low | Low | Low |
| Ramström, 1993 ^119^ | Low | Low | Low | Low | Low | Low |
| Jabalameli, 2006 ^64^ | Unclear | Unclear | Unclear | Unclear | Low | Low |
| Alimian, 2011 ^120^ | Low | Low | Low | Low | Low | Low |
| Eldaba, 2013 ^58^ | Low | Low | Unclear | Low | Low | Low |
| Langille, 2013 ^121^ | Low | Low | Low | Low | Low | Low |
| Jahanshahi, 2014 ^122^ | Low | Low | Low | Low | Low | Low |
| Shehata, 2014 ^42^ | Unclear | Low | Low | Low | Low | Unclear |
| El Shal, 2015 ^123^ | Low | Low | Low | Low | Low | Low |
| Nuhi, 2015 ^49^ | Low | Unclear | Low | Unclear | Low | High |
| Sakallioğlu, 2015^89^ | Low | Unclear | Unclear | Low | Unclear | Low |
| Eftekharian, 2016 ^82^ | Low | Low | Low | Low | Unclear | Low |
| Baradaranfar, 2017 ^44^ | Low | High | Low | Low | Low | High |
| Ghavimi, 2017 ^124^ | Low | Low | Low | Low | Low | Low |
| Dongare, 2018 ^46^ | Low | Unclear | Low | Low | Low | High |
| Quiroga, 2018 ^51^ | Low | High | Low | Low | Low | Low |
| Padhy, 2019 ^50^ | Low | Low | Unclear | High | Low | Low |
| Pannerselvam, 2019 ^88^ | Low | Low | Unclear | Unclear | Low | Low |
| Yang, 2021 ^125^ | Low | Low | Low | Low | Low | Low |
| Karaaslan, 2014 ^83^ | Low | Low | Low | Low | Unclear | Low |
| Karaaslan, 2015 ^126^ | Low | Low | Low | Low | Low | Low |
| Chiang, 2019 ^81^ | Low | Unclear | Low | Low | Low | Low |
| Felli, 2019 ^127^ | Low | Low | Low | Low | Low | Low |
| Pande, 2019 ^87^ | Unclear | Low | Low | Unclear | Low | Low |
| Lee, 2020 ^84^ | Low | Unclear | Low | Low | Low | Low |
| Fried, 2021 ^47^ | Low | Low | High | High | Low | Low |
| Ma, 2021 ^128^ | Low | Low | Low | Low | Low | Low |
| Liu, 2020 ^131^ | Low | Low | Low | Low | Low | Low |
| Bayram, 2021 ^129^ | Low | Low | Low | Low | Low | Low |
| Takahashi, 2021 ^90^ | Low | Low | Low | Low | Low | Unclear |
| Mackenzie, 2022 ^130^ | Low | Low | Low | Low | Low | Low |
| Nicholson, 2022 ^85^ | Low | Unclear | Low | Unclear | Low | Low |
| Nugent, 2019 ^86^ | Low | Low | Low | Unclear | Low | Low |

**Appendix 3**

1. **Investigation of publication bias**
1. Total volume of blood loss

| A. **Surgical field labelled** | B. **Route of administration labelled** |
| --- | --- |
| 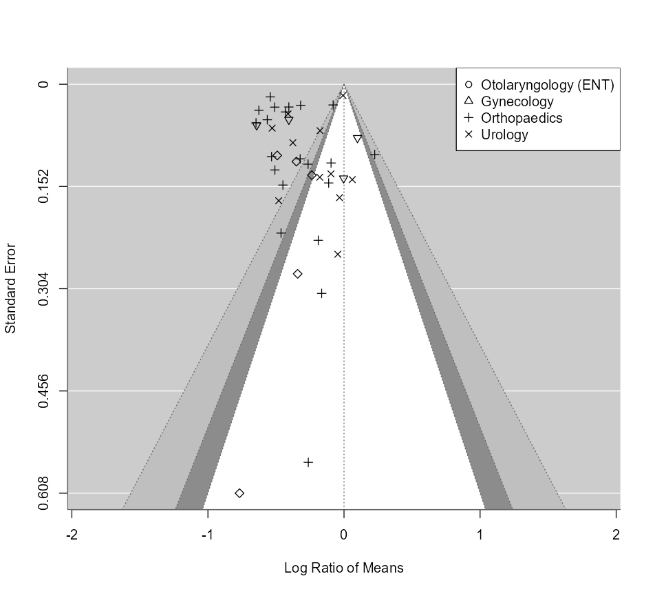 | 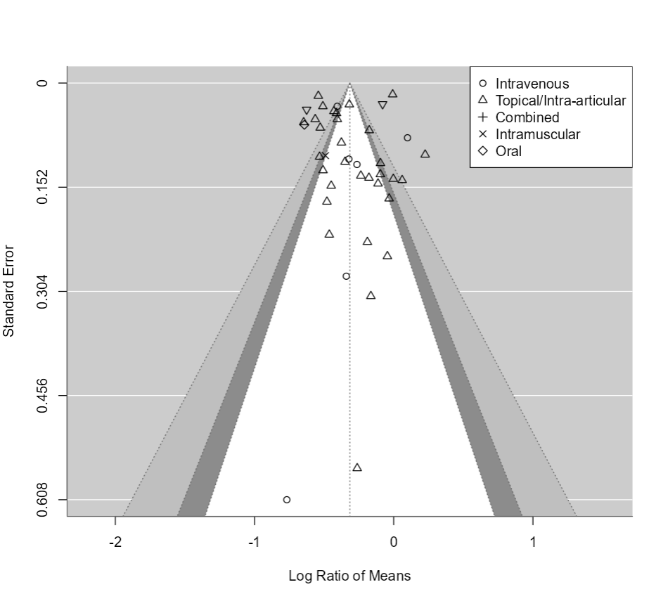 |

Figure 1: Contour Enhanced Funnel plots for log ratio of means of Total Blood Loss A: with Surgical Field Labelled B: with Route of Administration Labelled

2. Participants requiring blood transfusion

| A. | B. |
| --- | --- |
| 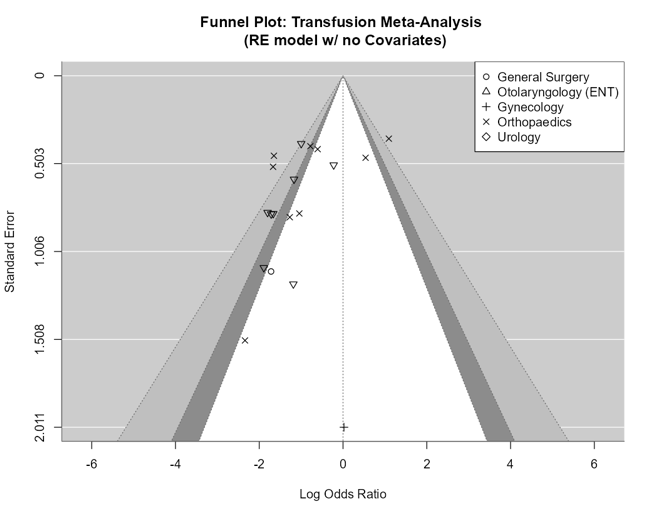 | 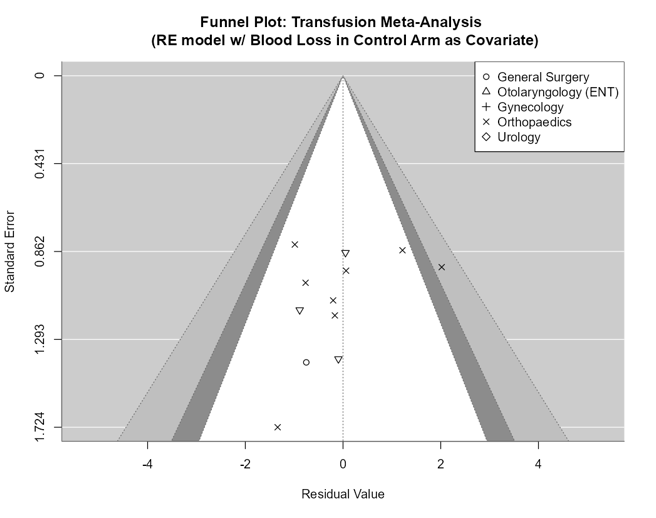 |

Figure 2: Contour enhanced funnel plots for log odds ratio of transfusion with A. No covariates in the model and B. Model adjusted for TBL in control arm as covariate

The funnel plot for the (log) odds ratio of transfusion has strong asymmetry which is mostly outside the 90% confidence contours suggesting studies with low statistical significance may be missing. However, when adjusting for blood loss in the control arm, the funnel plot of residual values from the fitted meta-analysis model produced a funnel plot with very low asymmetry.

2. **Subgroup analysis:**
   1. Total blood volume loss

| **A** | **B** |
| --- | --- |
| 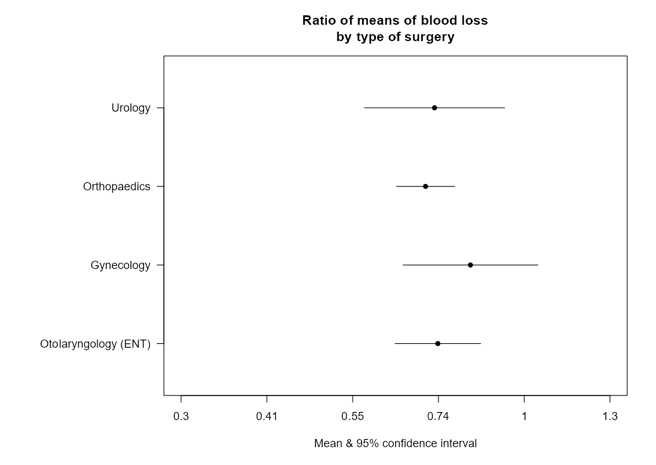 | 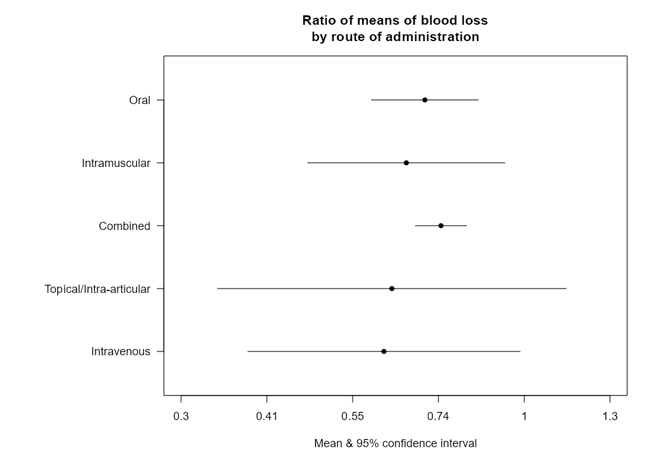 |

Figure 3: Subgroup analyses for A. Types of surgery and B. Route of TXA administration for the blood loss outcome.

2. Number of participants requiring blood transfusion:

| **A** | **B** |
| --- | --- |
| 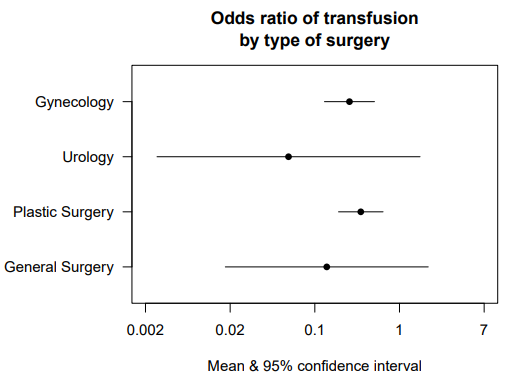 | 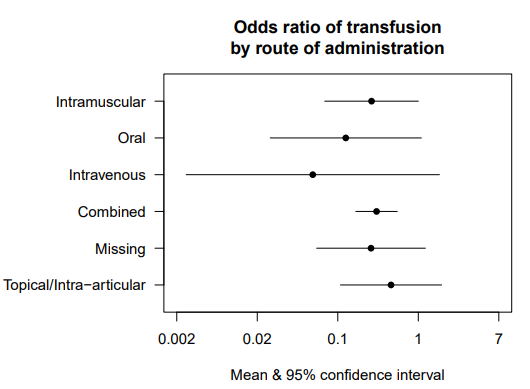 |

Figure 4: Subgroup analyses for odds ratio of transfusion by A. Surgical field and B. Route of TXA administration for the transfusion risk outcome

3. **Additional sensitivity analysis—Length of hospital stay**

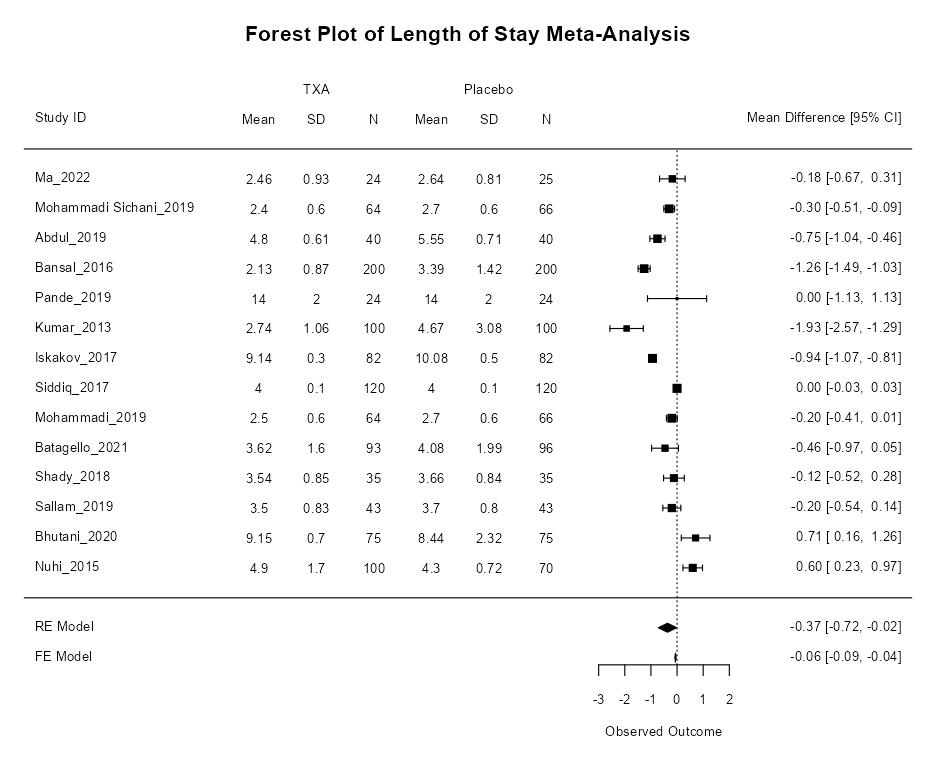

Figure 5: Additional sensitivity analysis for Length of hospital stay excluding the single influential trial

**Appendix 4**

**Exploratory analysis including the studies from orthopaedic speciality as an exemplar for surgeries at a high risk for blood loss**

A total of 91 RCTs (Table 1) with 8887 adult participants undergoing orthopaedic surgeries classified as moderate- to high-risk of blood loss were included. These RCTs were used to explore the effectiveness of TXA in preventing blood loss and need for transfusion in higher risk patients; specifically to facilitate the estimation of effectiveness response curves as a function of observed blood loss in trial control group.

1. Total Blood Loss

A meta-regression of the studies for mean difference of total blood loss (TBL), with TBL in the control arm as a covariate was performed. In the figure 1 black circles represent surgeries at low risk for blood loss while blue triangles represent surgeries at high risk of blood loss in this case from orthopaedics specialities. The regression slope has a gradient of –0.30 (95% CI –0.36, -0.23), which suggests for any increase in blood loss for a surgery, TXA reduces the blood loss by roughly one third i.e. this provides evidence that the difference in blood loss between groups is dependent on the blood loss in the control group. These figures, along with the plausible explanation that a surgery with higher TBL has more blood to “save”, led us to perform a meta-analysis of the ratio of means (RoM) for TBL since the absolute amount of blood “saved” by TXA is implicitly assumed to be a function of the blood lost in the control group for this outcome.

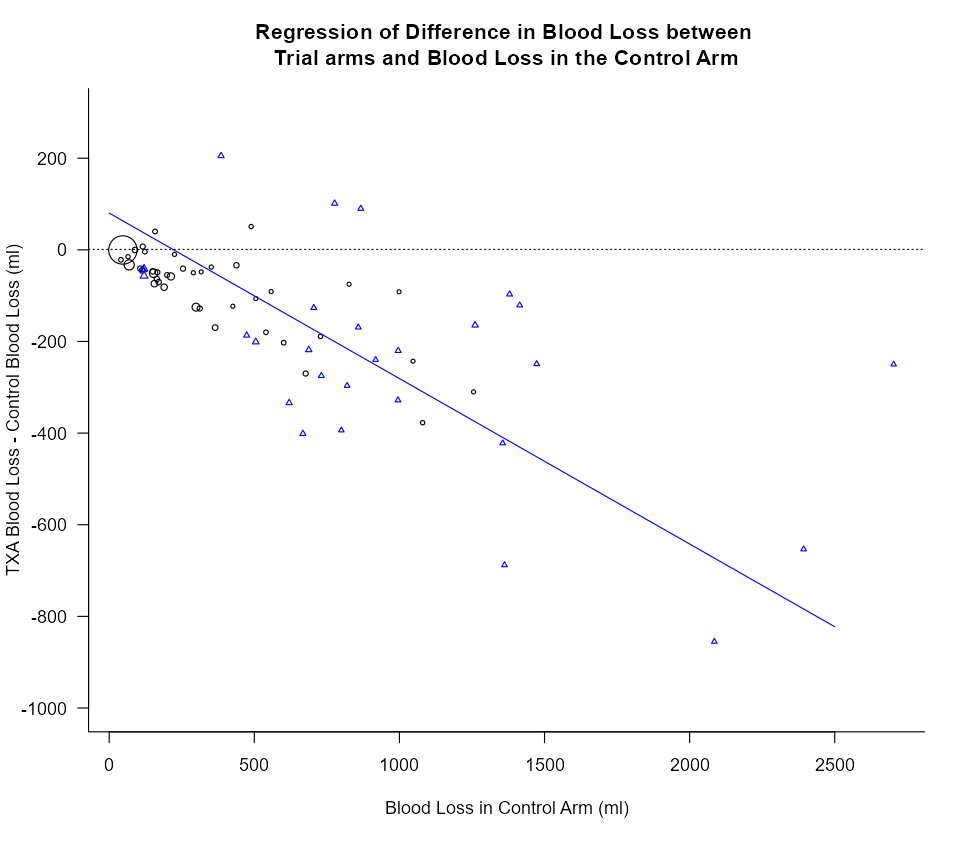

Figure 1: Scatter plot of mean difference in Total blood loss in TXA vs Total blood loss in control arm with regression line

| A. | B. |
| --- | --- |
| 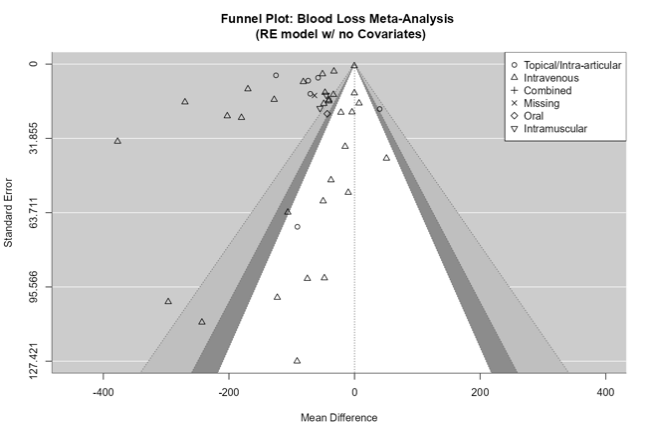 | 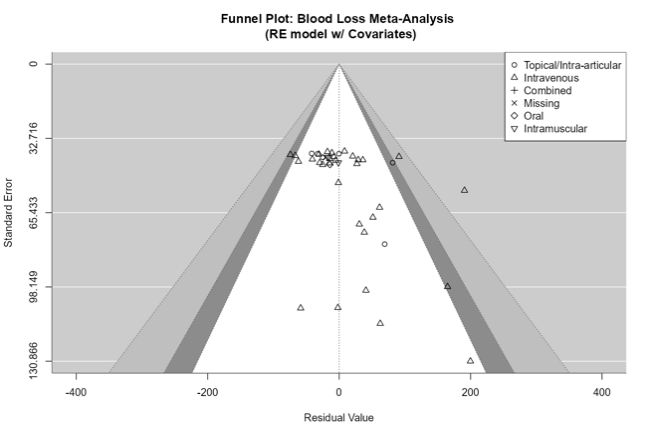 |

Figure 2: Contour enhanced funnel plots for Mean Difference of Total Blood Loss with A. No covariates in the model and B. Model adjusted for Total Blood Loss in control arm as covariate

2. Blood transfusion

A meta-regression of transfusion odds ratios with blood loss in the control arm as a covariate was conducted (Figure 2). The sizes of each trial on the plot also indicates the sample size of the trial, a bigger point means a bigger trial. Three regression slopes were plotted in the figure. The black regression slope representing low-risk trials only (black circles), with a slope of 0.0004 and a p-value of 0.6805. This means the odds ratio of transfusion for TXA increases by 0.04 (95% CI –0.17, 0.26) for every 100 ml increase of blood loss, which is not statistically significant. The blue regression slope representing the high-risk orthopaedics trials (blue triangles), suggests odds ratio of transfusion decreases by 0.09 (95% CI –0.30, 0.12) for every 100ml increase and is not statistically significant (p=0.3999). The final red regression slope is representing all trials combined, it indicates a 0.03 (95% CI –0.10, 0.17) increase in odds ratio of transfusion for every 100ml increase of blood loss, and is not statistically significant (p=0.6324). The three trials with blood loss larger than 2000ml were excluded from all regression analyses due to their extreme nature. These analyses concluded that there was little evidence that the odds ratio for requiring a transfusion varies systematically with the amount of blood loss over a large range of observed blood losses.

Although trials represented as black circles are classified as low-risk and those as blue triangles as medium- and high-risk, there is very large overlap in the amount of blood lost in the control arm of the trials. Also, only a small number of the triangles report blood loss in the control group of <500 ml (and those that do are close to this threshold), however, a large proportion of the black circles (low risk trials) report blood loss of over 500ml, some markedly over. This highlights variability in observed blood loss within risk classifications and highlights the challenges of predicting blood loss prior to an operation. This point is considered further in the discussion of the paper.

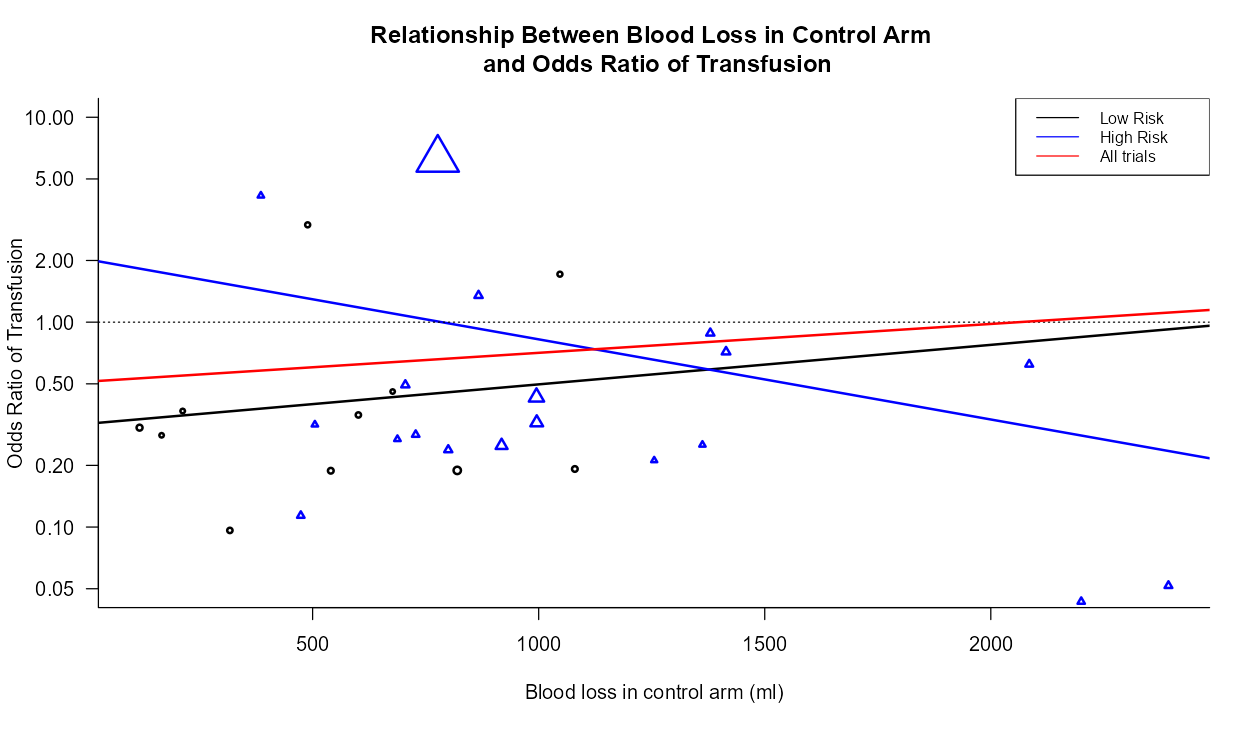

Figure 3: Meta regression of odds ratio of transfusion against TBL in control arm with regression slopes for different subgroups of trials

**Table 1: Orthopaedics studies classified as moderate to high risk for blood loss**

| Author | Year Published | Surgical area | No. of participants in Arm 1 | Arm 1 Intervention | No. of participants in Arm 2 | Arm 2 Intervention | No. of participants in Arm 3 | Arm 3 Intervention |
| --- | --- | --- | --- | --- | --- | --- | --- | --- |
| Cui XH ^1^ | 2015 | Knee Arthroplasty | 73 | Placebo | 73 | I/V and I/A TXA |  |  |
| Lin ^2^ | 2014 | Knee Arthroplasty | 40 | Placebo | 40 | I/V and I/A TXA |  |  |
| Neilipovitz ^3^ | 2001 | Spine | 18 | Placebo | 22 | I/V TXA |  |  |
| Tu ^4^ | 2015 | Knee Arthroplasty | 80 | Placebo | 66 | I/V and I/A TXA |  |  |
| Sethna ^5^ | 2005 | Spine | 21 | Placebo | 23 | I/V TXA |  |  |
| Xie ^6^ | 2015 | Hip Arthroplasty | 70 | I/V and I/A TXA | 70 | Topical TXA |  |  |
| Zhao Z ^7^ | 2015 | Knee Arthroplasty | 23 | Placebo | 24 | I/V and I/A TXA |  |  |
| Karaaslan ^8^ | 2014 | Knee Arthroplasty | 40 | Placebo | 41 | I/V and I/A TXA |  |  |
| Huang ZY ^9^ | 2014 | Knee Arthroplasty | 92 | I/V and I/A TXA | 43 | I/V TXA |  |  |
| Jain NP ^10^ | 2015 | Knee Arthroplasty | 60 | I/V TXA | 59 | I/V and I/A TXA |  |  |
| Zhao GH ^11^ | 2015 | Knee Arthroplasty | 70 | Placebo | 70 | I/A and Oral TXA |  |  |
| Karkar ^12^ | 2009 | Knee Arthroplasty | 13 | I/V Normal saline | 13 | I/V TXA |  |  |
| MacGillivray ^13^ | 2011 | Knee Arthroplasty | 20 | I/V Normal saline | 20 | I/V TXA |  |  |
| Kim ^14^ | 2014 | Knee Arthroplasty | 73 | I/V Normal saline | 73 | I/V TXA |  |  |
| Hafeez ^15^ | 2014 | Knee Arthroplasty | 15 | I/V Normal saline | 19 | I/V TXA |  |  |
| Shinde ^16^ | 2015 | Knee Arthroplasty | 14 | I/V Normal saline | 14 | I/V TXA |  |  |
| Chen ^17^ | 2016 | Knee Arthroplasty | 60 | I/V Normal saline | 60 | I/VTXA |  |  |
| Vara ^18^ | 2017 | Shoulder | 53 | I/V TXA | 49 | No TXA |  |  |
| Ersin ^19^ | 2020 | Shoulder | 32 | I/V TXA | 28 | Placebo |  |  |
| Bayram ^20^ | 2021 | Shoulder | 43 | I/A TXA | 47 | I/A Epinephrine in saline |  |  |
| Takahashi ^21^ | 2021 | Shoulder | 33 | I/V TXA | 33 | Placebo |  |  |
| Nicholson ^22^ | 2022 | Shoulder | 50 | I/V TXA | 50 | No TXA |  |  |
| Wang ^23^ | 2013 | Spine | 30 | I/V TXA | 30 | Placebo |  |  |
| Kim ^24^ | 2017 | Spine | 24 | I/V TXA | 24 | Placebo |  |  |
| Shi ^25^ | 2017 | Spine | 50 | I/V TXA | 46 | Placebo |  |  |
| Xu ^26^ | 2020 | Spine | 40 | I/V TXA | 40 | Placebo |  |  |
| Zhang ^27^ | 2020 | Spine | 138 | I/V TXA | 151 | Placebo |  |  |
| Zhu ^28^ | 2020 | Spine | 50 | I/V TXA | 50 | Placebo |  |  |
| Yan ^29^ | 2021 | Spine | 40 | I/V TXA | 40 | Placebo |  |  |
| Elwatidy ^30^ | 2008 | Spine | 32 | I/V TXA | 32 | Placebo |  |  |
| Sun ^31^ | 2019 | Spine | 26 | I/V TXA | 37 | Placebo |  |  |
| Li ^32^ | 2021 | Spine | 212 | I/V TXA | 227 | Placebo |  |  |
| Ou ^33^ | 2018 | Spine | 59 | I/V + I/A + Oral TXA | 59 | Placebo |  |  |
| Mu ^34^ | 2018 | Spine | 45 | I/V TXA | 42 | Placebo |  |  |
| He ^35^ | 2020 | Spine | 20 | I/V TXA | 20 | Placebo |  |  |
| Wong ^36^ | 2008 | Spine | 73 | I/V TXA | 74 | Placebo |  |  |
| Wang ^37^ | 2017 | Spine | 39 | I/A TXA | 41 | Placebo |  |  |
| Verma ^38^ | 2014 | Spine | 36 | I/V TXA | 47 | Placebo |  |  |
| Goobie ^39^ | 2018 | Spine | 56 | I/V TXA | 55 | Placebo |  |  |
| Liang ^40^ | 2016 | Spine | 30 | I/A TXA | 30 | Placebo |  |  |
| Xu ^41^ | 2017 | Spine | 40 | I/A TXA | 40 | Placebo |  |  |
| Hasan ^42^ | 2021 | Spine | 83 | I/V TXA | 83 | Placebo |  |  |
| Huang ^43^ | 2011 | Spine | 34 | I/V TXA | 34 | Placebo |  |  |
| Huang ^44^ | 2015 | Spine | 30 | I/V TXA | 30 | Placebo |  |  |
| Yan ^45^ | 2015 | Spine | 33 | Placebo | 35 | I/V TXA |  |  |
| Zhang ^46^ | 2015 | Spine | 38 | No TXA | 35 | I/V TXA |  |  |
| Feng ^47^ | 2016 | Spine | 60 | Placebo | 60 | I/V TXA |  |  |
| Jia ^48^ | 2016 | Spine | 30 | Placebo | 30 | I/V TXA |  |  |
| Nian ^49^ | 2016 | Spine | 30 | Placebo | 30 | I/A TXA |  |  |
| Meng ^50^ | 2017a | Spine | 40 | No TXA | 40 | I/A TXA |  |  |
| Meng ^50^ | 2017 b | Spine | 40 | No TXA | 40 | I/V TXA |  |  |
| Song ^51^ | 2017 | Spine | 16 | Placebo | 16 | I/V TXA |  |  |
| Zhang ^52^ | 2017 | Spine | 41 | Placebo | 41 | I/A TXA |  |  |
| Zhang ^52^ | 2017 | Spine | 41 | Placebo | 41 | I/V TXA |  |  |
| Chen ^53^ | 2018 | Spine | 100 | Placebo | 100 | I/V TXA |  |  |
| Hu ^54^ | 2018 | Spine | 40 | Placebo | 40 | I/V TXA |  |  |
| Wang ^37^ | 2018 | Spine | 41 | Placebo | 39 | I/V TXA |  |  |
| Zhang ^55^ | 2018 | Spine | 50 | Placebo | 54 | I/V TXA |  |  |
| Elmose ^56^ | 2019 | Spine | 116 | Placebo | 117 | I/V TXA |  |  |
| Liu ^57^ | 2019 | Spine | 35 | Placebo | 35 | I/V TXA |  |  |
| Wang ^58^ | 2019 | Spine | 28 | Placebo | 30 | I/V TXA |  |  |
| Xia ^59^ | 2020 | Spine | 44 | Placebo | 46 | I/A TXA |  |  |
| Yang ^60^ | 2019 | Spine | 16 | Placebo | 18 | I/V+ I/A+ Oral TXA |  |  |
| Zhu ^61^ | 2019 | Spine | 39 | Placebo | 39 | I/V TXA |  |  |
| Ding ^62^ | 2020 | Spine | 15 | Placebo | 15 | I/V TXA |  |  |
| Ding ^62^ | 2020 | Spine | 15 | Placebo | 15 | I/V TXA |  |  |
| Liu ^63^ | 2021 | Spine | 40 | Placebo | 40 | I/V TXA |  |  |
| Mi ^64^ | 2021 | Spine | 50 | Placebo | 50 | I/A TXA |  |  |
| Zhang ^65^ | 2021 | Spine | 40 | Placebo | 40 | I/V + I/A + Oral TXA |  |  |
| Farrokhi ^66^ | 2011 | Spine | 38 | Placebo | 38 | I/V TXA |  |  |
| Tsutsumimoto ^67^ | 2011 | Spine | 20 | Placebo | 20 | I/V TXA |  |  |
| Benoni ^68^ | 1996 | Knee Arthroplasty | 48 | Placebo | 48 | I/V TXA |  |  |
| Alvarez ^69^ | 2008 | Knee Arthroplasty | 55 | Placebo | 55 | I/V TXA |  |  |
| Alvarez ^70^ | 2019 | Knee Arthroplasty | 11 | Placebo | 11 | I/V TXA |  |  |
| Gautam ^71^ | 2013 | Knee Arthroplasty | 13 | No TXA | 14 | I/V TXA |  |  |
| Guerreiro ^72^ | 2017 | Knee ACL | 21 | No TXA | 22 | I/A TXA |  |  |
| Lee ^73^ | 2013 | Hip | 34 | Placebo | 34 | I/V TXA |  |  |
| Perez-Jimeno ^74^ | 2018 | Hip | 151 | No TXA | 142 | I/A TXA |  |  |
| Yue ^75^ | 2014 | Hip | 51 | Placebo | 52 | I/A TXA |  |  |
| Hiippala ^76^ | 1995 | Knee Arthroplasty | 13 | Placebo | 15 | I/V TXA |  |  |
| Hiippala ^77^ | 1997 | Knee Arthroplasty | 38 | Placebo | 39 | I/V TXA |  |  |
| Jansen ^78^ | 1999 | Knee Arthroplasty | 21 | Placebo | 21 | I/V TXA |  |  |
| Veien ^79^ | 2002 | Knee Arthroplasty | 15 | Placebo | 15 | I/V TXA |  |  |
| Good ^80^ | 2003 | Knee Arthroplasty | 27 | Placebo | 28 | I/V TXA |  |  |
| Orpen ^81^ | 2006 | Knee Arthroplasty | 15 | Placebo | 15 | I/V TXA |  |  |
| Painter ^82^ | 2018 | mixed hip and knee replacement | 69 | Placebo | 71 | I/V TXA |  |  |
| Yang ^83^ | 2020 | Knee Arthroplasty | 48 | Placebo | 48 | I/A TXA |  |  |
| Zhang ^84^ | 2007 | Knee Arthroplasty | 51 | Placebo | 51 | I/V TXA |  |  |
| Lin ^85^ | 2012 | Knee Arthroplasty | 50 | Placebo | 52 | I/V TXA | 49 | I/V TXA |
| Oztas ^86^ | 2015 | Knee Arthroplasty | 30 | No TXA | 30 | I/VTXA | 30 | I/A TXA |
| Clave ^87^ | 2019 | Hip | 75 | Placebo | 76 | I/V TXA (low dose) | 78 | I/V TXA (high dose) |
| Tanaka ^88^ | 2001 | Knee Arthroplasty | 26 | Placebo | 24 | I I/V TXA (low dose) | 22 | I/V TXA (high dose) |
| Camarasa ^89^ | 2006 | Knee Arthroplasty | 60 | Placebo | 35 | I/V TXA | 33 | I/V EACA |
| Stowers ^90^ | 2017 | Knee Arthroplasty | 30 | Placebo | 60 | I/A TXA | 60 | I/V TXA |
| Xue ^91^ | 2021 | Knee Arthroplasty | 53 | Placebo | 50 | I/V TXA (pre-op) | 53 | I/V TXA (pre- & intra- op) |

**Appendix 5**

**Cost-effectiveness Analysis**

**Introduction**

This appendix provides further information regarding the economic analyses presented in this paper. A decision analytic model was developed based on the modelling of interventions to reduce blood transfusion in surgical patients for NICE clinical guideline 24 (NG24). Analyses in NG24 addressed separate decision problems relating to surgeries with high (>1L), and moderate (0.5 - 1.0L) expected blood loss. NG24 addressed a wider set of potential interventions, including cell-salvage, which would not be considered in cases with lower expected volumes of blood loss. We therefore modified the model of NG24 to reflect the low expected blood loss setting. Informed by the results of the clinical review we excluded 30-day mortality. With no other 30-day outcome that would predict differing health or resource use beyond this point, our analysis is conducted over the short term of the surgical admission. Rather than model the quantity of blood transfused as was done for higher expected blood loss in NG24, we made the simplifying assumption, based on clinical advice, that use of more than one unit of transfused blood would not be required. These modifications to the NG24 model structure mean the analyses are driven by predicted reductions in transfusions and length of stay, with a quality-of-life distinction applied to the latter.

As in NG24 we constructed the model probabilistically (with 10,000 iterations) with probability distributions defined for relevant input parameters. Both the base case and scenario analyses are reported on the basis of probabilistic analyses. In the absence of any mortality effect quality of life gains are minor, and rather than present incremental cost-effectiveness ratios (ICERs), we present expected incremental net benefits.

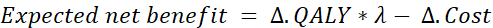

where
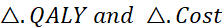
 are the incremental total QALYs and costs of TXA versus standard care, and lambda is the cost-effectiveness threshold. We hold lambda constant at £20,000 per QALY.

As the acquisition cost of TXA is low relative to the cost of blood transfusions, a very simple calculation can indicate the small probability of blood transfusion that would be required for TXA to be cost saving assuming other factors remain constant, irrespective of how low the transfusion risk were to fall. We quantify the expected net benefit across levels of transfusion risk. Probabilistic one-way sensitivity analyses for key individual parameters are presented as described by McCabe et al^133^. Distributions for clinical parameters are based on the findings of the clinical review and meta-analysis. We extended uncertainty analyses to include lower transfusion probabilities in patients not treated with TXA those resulting from the meta-analysis. We did not assign a probability distribution to either the cost of TXA or blood transfusion but did assume a standard error for the cost of a hospital ward day equal to 15% of the mean.

**Base case analysis**

**Model parameters**

Model parameters are the probability of transfusion, the effect of TXA on need for transfusion (log-odds ratio), effect on length of stay, unit costs (TXA, transfusion, hospital bed per diem), and the impact in terms of health-related quality of life (utility) associated with being in hospital. We applied results from the meta-analyses based on random effects. These base case parameters are summarized in Table 1.

Table 1. Model parameters for the base case analysis

| **Parameter** | **Estimate (95% confidence interval / S.E.) [distribution]** | **Source** |
| --- | --- | --- |
| Log odds transfusion (untreated) | -1.605 (-1.995, -1.214) [log odds] | Meta-analysis |
| Log odds ratio transfusion TXA | -0.945 (-1.404, -0.487) [log odds ratio] | Meta-analysis |
| Difference in length of stay (days) | 0.397 (0.020, 0.774) [normal] | Meta-analysis |
| Costs |  |  |
| TXA (per patient) | £1.66 | eMIT (2024) |
| Unit blood transfused | £216.53 | NHS (2024) |
| Hospital per diem | £345.00 (51.75) [gamma] | Hansard (2023) |
| Utility |  |  |
| Hospitalized (per diem) | 0.247 (0.008) [normal] | NG24 (HSE, 2012) |

The clinical meta-analysis provided the pooled risk for transfusion using a random effects model. We assumed that the effect of TXA on length of stay was independent of the degree of blood loss in the control arms of the included studies and applied the overall random effects mean length of stay reduction.

The cost per surgery for TXA was calculated based on the weighted average price of TXA 1000mg/10ml solution for injection ampoules, which is £4.15 for a pack of 5 ampoules (£0.83 per ampoule). Given a typical dose of 2g administered via slow IV injection, the cost per surgery for TXA was determined as follows: £0.83 × 2 = £1.66. For blood transfusions, in addition to assuming a single unit of blood per transfusion, we also maintained the simplifying assumption of NG24 that all transfusions are of red blood cells (RBC), which would constitute most blood products transfused. The breakdown for the cost of blood transfusion, based on NG24, is reported in Table 2. For all surgical types we assumed common per diem cost for hospital stay. We retained the decrement for quality of life associated with a day in hospital applied in NG24.

**Results**

The results for our base case analysis are presented in Table 3. The expected rate of transfusion in the overall set of studies classified as low bleeding risk in our base case is 17%. With odds ratio of 0.35 this is reduced to 7% which, given our assumption of a single unit of red blood per transfusion, represents a saving of 0.095 units of blood per patient. A reduction in length of stay of an average 0.40 days would provide substantial additional cost-savings, which we also model as providing a small QALY gain. In total TXA is modelled in the base case to save £156 (95%CI; £25, £305) per patient, while producing a slight QALY gain and therefore dominating standard care. Net monetary benefit is £161, with a probability of being cost-effective of 0.99 at a cost-effectiveness threshold of £20,000 per QALY. Figure 1 illustrates the extent of dominance of TXA in the probabilistic analysis. The probability that TXA is resource saving in terms of blood transfusion is 1.00, and the 0.01 probability that TXA would not be cost-effective is due to there being a slight chance, based on the meta-analysis of length of stay, that the mean difference could narrowly favour standard care.

Table 2. Cost per blood transfusion

| **Component** | **Mean time (min)** | **Staff cost per min (£)** | **Mean cost (£)** | **Assumptions & sources** |
| --- | --- | --- | --- | --- |
| **Staff time (blood bank)** |  |  |  |  |
| Computer issue | 5.38 | £0.83 | £4.48 | Staff time from Agrawal 2006 Staff unit costs: Unit Costs of Health and Social Care 2023 Table 11.1.2: Hospital-based scientific and professional staff band 4 and band 6 |
| Blood collection | 5 | £0.57 | £2.83 |  |
| Blood ordering | 1.02 | £0.83 | £0.85 |  |
| Blood delivery | 10 | £0.57 | £5.67 |  |
| **Staff time (ward)** |  |  |  |  |
| Collection and administration | 15 | £0.80 | £12.00 | Staff time based on NG24 GDG expert opinion. Staff unit costs: Unit Costs of Health and Social Care 2023 Table 11.2.2: Hospital-based nurses, qualified, band 5 |
| Observations | 25 | £0.80 | £20.00 |  |
| **Disposables (ward)** |  |  |  |  |
| Patient assessment |  |  | £3.09 | Agrawal (2006); £ values inflated to 2023-2024 |
| Transfusion preparation |  |  | £1.48 |  |
| Transfusion for 1st unit |  |  | £5.57 |  |
| **Blood product** |  |  |  |  |
| RBC per unit |  |  | £158.18 | NHS Blood and Transplant. Price list 2014/2015. £ values inflated to 2023-2024. NHS Blood and Transplant; 2014. Assumed all transfusion RBC. |
| Wastage per unit |  |  | £2.37 | Wastage assumed to be equal to 1.5% of the cost of a unit of RBC, based on reported rate from Agrawal (2006) |
| **Total cost** |  |  | **£216.53** |  |

Table 3. Base case analysis

|  | **SOC** | **TXA** | **Incremental** |
| --- | --- | --- | --- |
| **Outcomes** |  |  |  |
| Transfusions | 0.167 | 0.072 | 0.095 (0.05, 0.14) |
| Length of stay | 0.398 | 0 | 0.398 (0.02, 0.78) |
| QALY gain | 0.000 | 0.000 | 0.000 |
| **Costs (£)** |  |  |  |
| TXA | 0 | 1.66 | 1.66 |
| Transfusion | 36.06 | 15.59 | -20.47 (-30, -12) |
| Length of stay | 137.14 | 0 | -137.14 (-6, -285) |
| Total | 173.20 | 17.25 | -155.95 (-25, -305) |
| **Cost-effectiveness** |  |  |  |
| Cost per QALY | | TXA Dominant | |
| Expected NMB (£) at £20K / QALY | | | 161 (25, 315) |
| Probability cost-effective | |  | 0.990 |

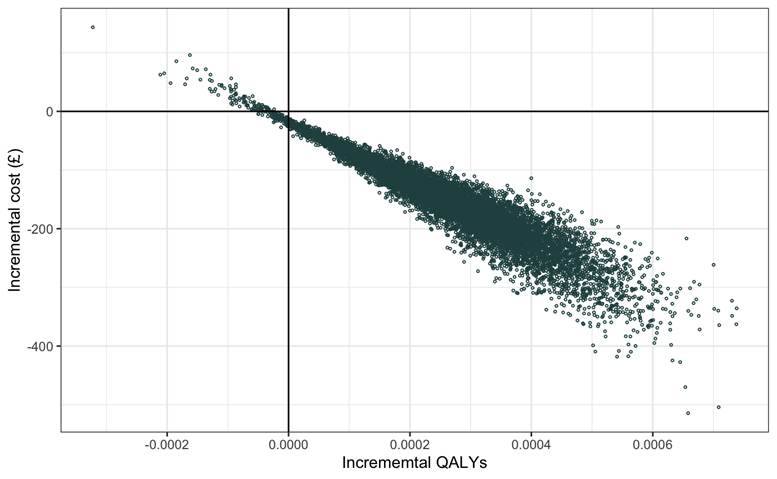

Figure 1. Cost-effectiveness plane base-case analysis

**Sensitivity analysis**

We varied the log odds and log odds ratio for transfusion, and the length of stay reduction, across quantiles of their respective distributions. We also performed analyses at levels of transfusion risk less than or equal to 10% to assess how net benefit would be impacted by lower levels of risk than seen in the clinical review.

With length of stay applied based on clinical review, the analysis was insensitive to variation over both the log odds and log odds ratio for transfusion (Figure 2A). Even without a length of stay effect, net benefit remains positive due to savings in blood transfusion (Figure 2B). Cost savings associated with length of stay are modelled independently of transfusion. Therefore, even as transfusion risk was assumed to be very low, net benefit remained relatively unaffected (Figure 3A). With length of stay effects excluded, net benefit is due solely to savings in blood transfusions. These savings remained positive other than at levels of blood transfusion below 2% (as a simple calculation would predict) (Figure 3B).

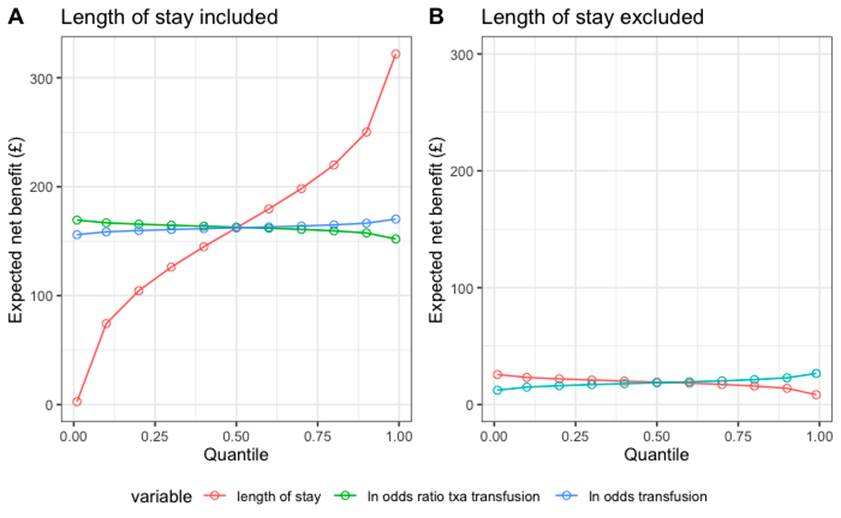

Figure 2. Sensitivity analyses for key parameters

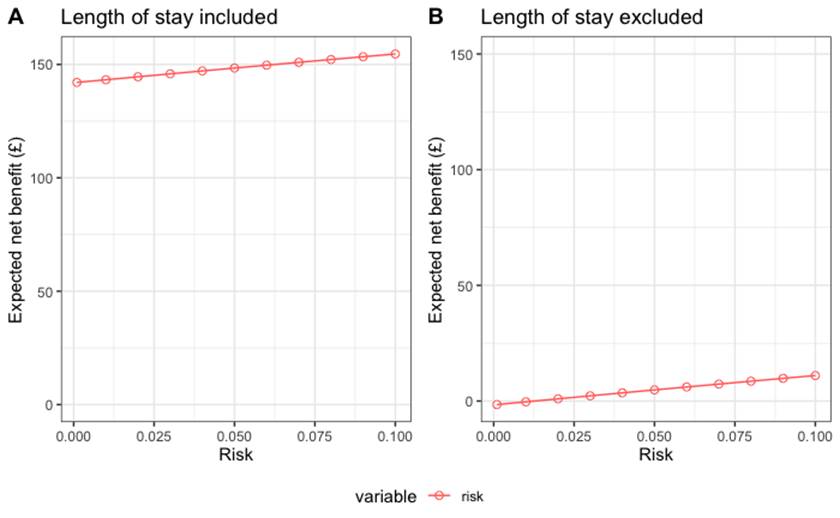

Figure 3. Sensitivity analyses at very low transfusion risk

**Scenario analysis**

In scenario analyses, we applied results from the meta-analyses based on fixed effects for the probability of transfusion, the effect of TXA on need for transfusion (log-odds ratio), and the effect on length of stay. These are summarized in Table 4.

Table 4: Model parameters for the scenario analysis

| **Parameter** | **Estimate (95% confidence interval / S.E.) [distribution]** | **Source** |
| --- | --- | --- |
| Log odds transfusion (untreated) | -1.468 (-1.619, -1.318) [log odds] | Meta-analysis |
| Log odds ratio transfusion TXA | -0.700 (-0.959, -0.441) [log odds ratio] | Meta-analysis |
| Difference in length of stay (days) | 0.601 (0.526, 0.676) [normal] | Meta-analysis |
| Surgery type |  |  |
| General surgery | -1.946 (-2.883, -1.009) [log odds] | Meta-analysis |
| Gynaecology | -1.160 (-1.851, -0.470) [log odds] | Meta-analysis |
| Otolaryngology | -4.500 (-7.287, -1.713) [log odds] | Meta-analysis |
| Plastic surgery | -1.291 (-2.235, -0.347) [log odds] | Meta-analysis |
| Urology | -2.013 (-2.237, -1.789) [log odds] | Meta-analysis |

Neither fixed effect estimates for standard care probability of transfusion nor the odds ratio for transfusion had any notable impact on cost-savings (Table 5). Applying the fixed effects estimate for mean difference in length of stay notably increased the saving, which remained the case when fixed effects estimates were simultaneously applied for baseline transfusion risk and log odds ratio for TXA.

We found no modifying effect of surgery type on the odds ratio for transfusion with TXA. There are some suggestions, however, of differences in baseline transfusion risk across surgery types. Sensitivity analysis above shows TXA remaining cost-effective at all levels of baseline risk but depending increasingly on length of stay as baseline risk declines. For general surgery and urology, the mean risk was approximately 12%-14%, based on meta-analyses of nine and eight studies respectively. The three other surgical types are represented by only a single study each. For the gynaecology and plastic surgery studies, baseline transfusion risk was notably higher, at approximately 25%, but with substantially overlapping 95% confidence intervals across these categories. The one notably lower risk category was otolaryngology. The mean risk included in the meta-analysis in this category is similar to the lower limit for baseline risk suggested by the sensitivity analysis addressing baseline risk below 10%. In fact, in the single otolaryngology study included here, there were zero transfusions, and the risk is derived from a continuity correction. Nevertheless, we include this group as an illustration of a particularly low-risk surgical category.

Transfusions were avoided in fewer than 1% of cases in the otolaryngology group. The probability of TXA being cost-effective at a threshold of £20,000 per QALY remained approximately 0.98 due to the length of stay savings (Table 6). In this surgery type when difference in length of stay is excluded, there are limited savings due to transfusions avoided resulting in the greatly reduced probability that TXA would be cost-effective.

Table 5. Scenario analyses - fixed effects: incremental costs and net monetary benefit

|  | **Transfusions**  **(95% CI)** | **Transfusion Cost (£) (95% CI)** | **LOS Cost (£) (95% CI)** | **Total Cost (£) (95%CI) NMB (pCE)** | **Total Cost (£) excl. LOS (95%CI) NMB (pCE)** |
| --- | --- | --- | --- | --- | --- |
| Base case | 0.09 (0.05, 0.14) | -20 (-30, -12) | -137 (-6, -285) | -156 (-25, -305) | -19 (-10, -28) |
|  |  |  |  | 161 (0.990) | 19 (1) |
| Transfusion log odds fixed effects | 0.1 (0.06, 0.14) | -22 (-30, -14) | -137 (-6, -285) | -158 (-27, -307) | -21 (-12, -28) |
|  |  |  |  | 164 (0.991) | 21 (1) |
| Transfusion log OR fixed effects | 0.08 (0.05, 0.11) | -17 (-24, -11) | -137 (-6, -285) | -152 (-22, -301) | -15 (-9, -22) |
|  |  |  |  | 159 (0.989) | 15 (1) |
| LOS fixed effects | 0.09 (0.05, 0.13) | -20 (-29, -11) | -207 (-148, -279) | -225 (-165, -298) | -18 (-10, -27) |
|  |  |  |  | 234 (1) | 18 (1) |
| All fixed effects | 0.08 (0.06, 0.11) | -18 (-24, -13) | -207 (-148, -279) | -224 (-164, -295) | -17 (-11, -22) |
|  |  |  |  | 232 (1) | 17 (1) |

Table 6. Scenario analyses – surgery type: incremental costs and net monetary benefit

|  | **Transfusions**  **(95% CI)** | **Transfusion Cost (£) (95% CI)** | **LOS Cost (£), (95% CI)** | **Total Cost (£), (95%CI) NMB (pCE)** | **Total Cost (£) excl. LOS (95%CI) NMB (pCE)** |
| --- | --- | --- | --- | --- | --- |
| General surgery | 0.07 (0.03, 0.14) | -15 (-31, -6) | -137 (6, -285) | -151 (-19, -299) | -14 (-4, -29) |
|  |  |  |  | 156 (0.988) | 14 (1) |
| Gynaecology | 0.13 (0.06, 0.21) | -27 (-45, -14) | -137 (6, -285) | -163 (-31, -313) | -26 (-12, -43) |
|  |  |  |  | 168 (0.992) | 26 (1) |
| Otolaryngology | 0.01 (0, 0.04) | -1 (-8, 0) | -137 (6, -285) | -137 (-6, -285) | 0 (2, -7) |
|  |  |  |  | 142 (0.981) | 0 (0.209) |
| Plastic surgery | 0.11 (0.05, 0.21) | -24 (-45, -10) | -137 (6, -285) | -160 (-29, -308) | -23 (-8, -43) |
|  |  |  |  | 165 (0.991) | 23 (1) |
| Urology | 0.07 (0.04, 0.09) | -15 (-20, -9) | -137 (6, -285) | -156 (-20, -299) | -13 (-7, -18) |
|  |  |  |  | 156 (0.989) | 13 (1) |

**Appendix 6**

**Deviations from Protocol:**

a. Outcomes—We had planned to analyse only number of transfusions as primary outcome, but we included additional primary outcomes – total blood volume loss and the safety outcome - Deep vein thrombosis
b. Analysis—we had planned to analyse the mean difference for total blood volume loss, but we used ratio of mean volume loss as the primary analysis due to it fitting the data better.
c. We did not analyse outcomes—Mortality, Quality of life, Blood volume transfused, Surgical bleeding, Post operative bleeding, Adverse events: Acute Myocardial infarction; postoperative thrombosis and rate of serious adverse event due to unavailability of such data from published papers and reports
